# Target-agnostic drug prediction integrated with medical record analysis uncovers differential associations of statins with increased survival in COVID-19 patients

**DOI:** 10.1101/2022.04.12.22273802

**Authors:** Megan M. Sperry, Tomiko Oskotsky, Ivana Marić, Shruti Kaushal, Takako Takeda, Viktor Horvath, Rani K. Powers, Melissa Rodas, Brooke Furlong, Mercy Soong, Pranav Prabhala, Girija Goyal, Kenneth E. Carlson, Ronald J. Wong, Idit Kosti, Brian L. Le, James Logue, Holly Hammond, Matthew Frieman, David K. Stevenson, Donald E. Ingber, Marina Sirota, Richard Novak

## Abstract

**Importance:** Drug repurposing requires distinguishing established drug class targets from novel molecule-specific mechanisms and rapidly derisking their therapeutic potential in a time-critical manner, particularly in a pandemic scenario. In response to the challenge to rapidly identify treatment options for COVID-19, several studies reported that statins, as a drug class, reduce mortality in these patients. However, it is unknown if different statins exhibit consistent function or may have varying therapeutic benefit.

**Objectives:** To test if different statins differ in their ability to exert protective effects based on molecular computational predictions and electronic medical record analysis.

**Main Outcomes and Measures:** A Bayesian network tool was used to predict drugs that shift the host transcriptomic response to SARS-CoV-2 infection towards a healthy state. Drugs were predicted using 14 RNA-sequencing datasets from 72 autopsy tissues and 465 COVID-19 patient samples or from cultured human cells and organoids infected with SARS-CoV-2, with a total of 2,436 drugs investigated. Top drug predictions included statins, which were then assessed using electronic medical records containing over 4,000 COVID-19 patients on statins to determine mortality risk in patients prescribed specific statins versus untreated matched controls. The same drugs were tested in Vero E6 cells infected with SARS-CoV-2 and human endothelial cells infected with a related OC43 coronavirus.

**Results:** Simvastatin was among the most highly predicted compounds (14/14 datasets) and five other statins, including atorvastatin, were predicted to be active in > 50% of analyses. Analysis of the clinical database revealed that reduced mortality risk was only observed in COVID-19 patients prescribed a subset of statins, including simvastatin and atorvastatin. *In vitro* testing of SARS-CoV-2 infected cells revealed simvastatin to be a potent direct inhibitor whereas most other statins were less effective. Simvastatin also inhibited OC43 infection and reduced cytokine production in endothelial cells.

**Conclusions and Relevance:** Different statins may differ in their ability to sustain the lives of COVID-19 patients despite having a shared drug target and lipid-modifying mechanism of action. These findings highlight the value of target-agnostic drug prediction coupled with patient databases to identify and clinically evaluate non-obvious mechanisms and derisk and accelerate drug repurposing opportunities.

## Introduction

The emergence of the COVID-19 pandemic presented an urgent need for new and effective therapeutics, and repurposing of approved drugs with known safety profiles offered a path to identify viable treatment options. Research programs that focused on drug repurposing for COVID-19 presented opportunities to broadly uncover and understand new features of existing drugs.^1–3^ Recent retrospective studies by members of our group and others have shown that COVID-19 patients taking drugs from one of the most prescribed drug classes in the world – statins – exhibit a reduced mortality rate, but these studies pooled all statin compounds (e.g., lovastatin, simvastatin, atorvastatin, etc.) together in their analyses.^4–6^ All statins are prescribed to lower lipid and cholesterol levels, and share a common mechanism involving inhibition of HMG-CoA reductase (HMGCR); however, statins are also known to have anti-inflammatory and immunomodulatory properties, through mechanisms that involve several pathways,^5–11^ potentially by upregulating heme oxygenase-1 (HO-1).^7^ In addition, while three retrospective studies that pooled all statins demonstrated a significant reduction in mortality risk, no improvement in mortality outcome could be detected in another study.^10^ This raises the possibility that different statins might differ in their ability to reduce morbidity and mortality in COVID-19 patients, which could influence the results of studies based on which drugs were included. Moreover, if true, it would be important to distribute this information widely because it could influence clinical decision-making with regard to statin selection during the current COVID-19 crisis.^12,13^

Throughout the pandemic, multiple scientific teams predicted that existing drugs could be repurposed as potential COVID-19 therapeutics computationally through the application of high-throughput *in silico* screens based on artificial intelligence, network diffusion, or network proximity algorithms using the human interactome, SARS-CoV-2 targets, drug targets, docking structures, or biomedical literature as algorithmic inputs.^1–3^ These screens proposed hundreds of potential therapeutic options and led to further testing in SARS-CoV-2 infected culture systems and animal models.^1^ However, while *in vitro* and pre-clinical testing have offered promising predictions, clinical validation and translation of predicted compounds are much more challenging and few, if any, of these drugs proposed to be repurposed for COVID-19 have demonstrated clinical efficacy. Thus, there is a need for combining improved drug prediction capabilities, despite complex and often inadequately understood biology, with real world evidence, such as electronic health records (EHRs),^4,14^ to better inform which predicted compounds should advance toward clinical evaluation. A related approach has previously been used to identify repurposed drugs for coronary artery disease by integration of protein-protein interaction network proximity and large-scale patient-level longitudinal data.^15^

With drug repurposing in mind, we used a Network Model for Causality-Aware Discovery (NeMoCAD) computational tool based on Bayesian statistical network modeling^16^ to analyze transcriptomics signatures in tissue samples obtained from COVID-19 positive patients or SARS-CoV-2 infected human cell or organoid cultures to identify FDA-approved drugs that shift the host transcriptomic response to SARS-CoV-2 towards a healthy state (**Figure 1A**). This approach was agnostic as it was accomplished without an *a priori* defined drug target or mechanism of action.

**Figure 1.**
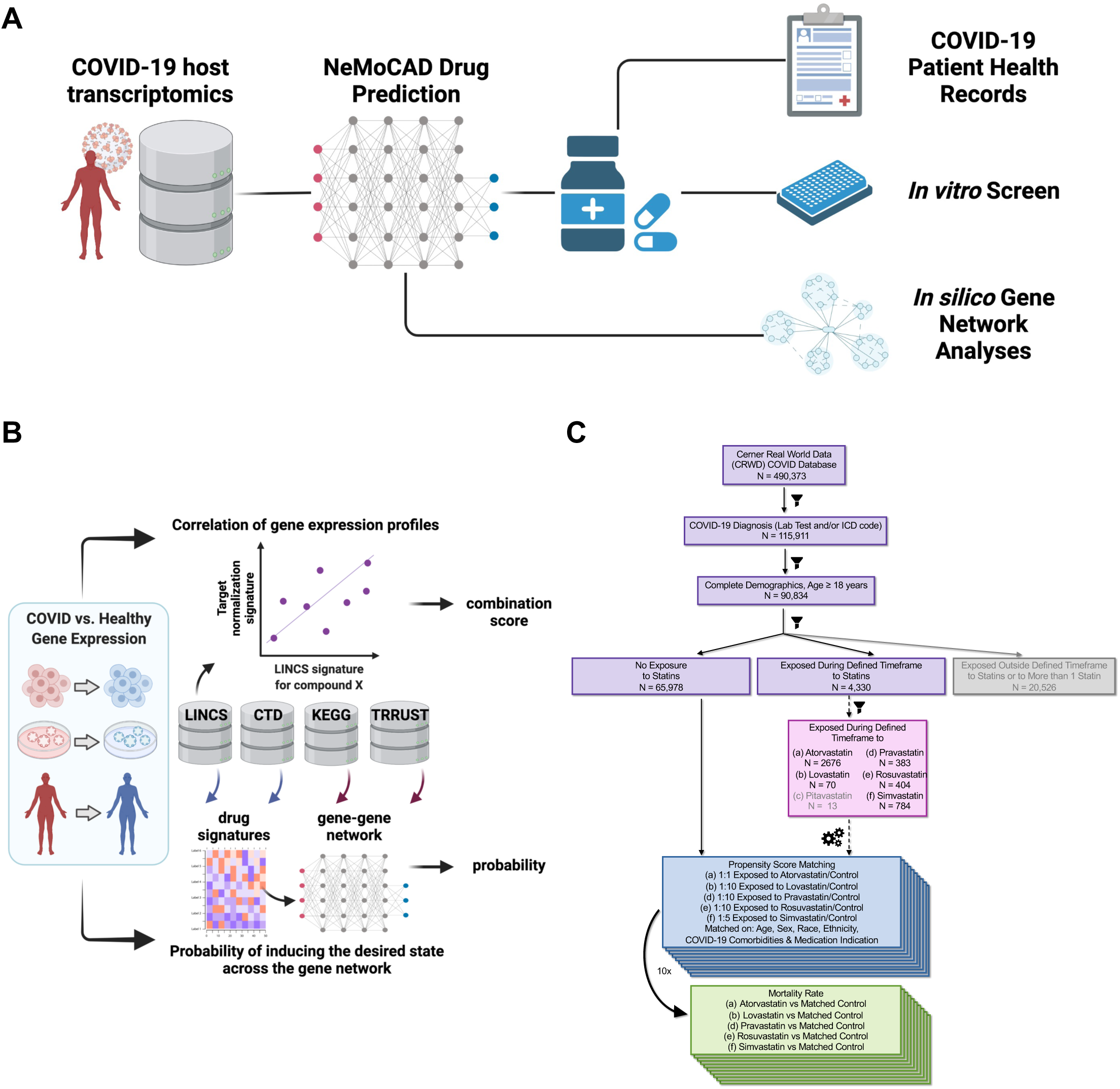
Combination approach for pathway-agnostic identification of compounds for drug repositioning. **(a)** The NeMoCAD gene network analysis tool is a drug repurposing algorithm that uses Bayesian statistical network analysis combined with data from publicly available datasets (e.g., LINCS, KEGG, CTD, TRRUST) for reference transcriptional signatures and to define regulatory network architecture. The algorithm identifies transcriptome-wide differential expression profiles between two biological states (e.g., healthy vs. diseased) in experimental or published transcriptomic datasets and defines the target normalization signature, i.e., the subset of genes that would need to reverse their expression to revert one state to the other. The output of NeMoCAD includes correlation and causation predictions for numerous chemical compounds and approved drugs in the LINCS database based on their ability to reverse the differential expression profile of interest. **(b)** Patient selection and analysis of electronic health records.

This computational analysis revealed that a subset of commonly administered statins were among the drugs most frequently predicted to revert the genome-wide gene expression profile of COVID-19 samples to that of a healthy state. Despite limited chemical diversity, statins induce a range of side effects that differ between individual statins^17,18^ suggesting the potential for distinct biological activities outside their known shared HMGCR mechanism, which potentially could be harnessed for drug repurposing and eventual new drug development. To explore this possibility in a clinical setting, a retrospective analysis was carried out using a database containing EHRs of over 490,000 COVID-19 patients, more than 4,000 of which are from patients who are actively taking statins (**Figure 1B**). This analysis demonstrated that use of only a subset of statins, including simvastatin and atorvastatin, correlated with decreased morbidity and increased survival in COVID-19 patients, confirming hidden divergent activities within a seemingly homogeneous drug class. Experimental *in vitro* studies confirmed that the drug most frequently predicted to reverse the COVID-19 state and that correlated with decreased morbidity in EHRs – simvastatin – also potently inhibited infection of Vero6 cells by SARS-CoV2 *in vitro*. However, other statins were less effective, suggesting that individual statins might have molecule-specific activities beyond the shared HMGCR target and thus vary in their ability to protect COVID-19 patients. ^14^

## Methods

### Transcriptomic-based compound prediction for drug repurposing

The drug prediction software, NeMoCAD (Network Modeling for Causal Discovery), was used to predict compounds that would mimic the shift from a COVID-19-positive state to a control state.^16^ NeMoCAD is a drug repurposing algorithm that performs correlation analysis of transcriptional gene signatures and a Bayesian statistical analysis of a network comprised of drug-gene and drug-drug interactions to identify compounds capable of changing a transcriptional signature indicative of disease to a healthy state.^16^ Using 14 publicly available transcriptomic datasets derived from human patients, tissue samples, organoids, and cells (**Table 1**),^19–25^ NeMoCAD identified transcriptome-wide differential expression profiles between the control and COVID-19 states for each dataset and defined a target normalization signature to mimic, which would shift the transcriptome from a COVID-19 disease to control state (**Supplemental Methods**). To understand underlying differences in LINCS drug-gene probability signatures that could influence drug predictions, statins were compared by principal component analysis (PCA). Using the time-averaged drug-gene probabilities from LINCS for 12,328 genes across 7 statins of interest (atorvastatin, fluvastatin, lovastatin, pitavastatin, pravastatin, rosuvastatin, and simvastatin), we created a PCA object in the R package ggfortify using the autoplot function. The package ggplot2 was used for plotting customizations. R version 4.0.5 was used for all computations and plotting.

**Table 1.** RNA-sequencing datasets used as inputs for network-based drug predictions.

| Dataset Source | Public Identifier/Link | Sample Source | Tissue/Cell Type | Conditions assessed for predictions | COVID-19 samples | Control samples | Drugs predicted |
| --- | --- | --- | --- | --- | --- | --- | --- |
| **Greninger<sup>17</sup>** | GSE152075 | Nasal swab samples from COVID patients | Nasopharyngeal | COVID > healthy | 430 | 54 | 377 |
| **Mertz<sup>18</sup>** | GSE151764 | Autopsy samples from COVID patients | Lung | COVID > healthy | 16 | 6 | 623 |
| **Mertz<sup>18</sup>** | GSE151764 | Autopsy samples from COVID patients | Lung | COVID (patients w/ normal BMI) > healthy | 2 | 6 | 647 |
| **Mertz<sup>18</sup>** | GSE151764 | Autopsy samples from COVID patients | Lung | COVID (patients w/ high BMI) > healthy | 14 | 6 | 571 |
| **OSF** | https://osf.io/7nrd3/ | COVID patients | Blood PBMC | COVID > healthy | 3 | 3 | 330 |
| **OSF** | https://osf.io/7nrd3/ | COVID patients | Bronchoalveolar lavage fluid | COVID > healthy | 32 | 54 | 436 |
| **Redmond<sup>16</sup>** | GSE151803 | Organoids | Liver | SARS-CoV-2 infected > mock treated | 6 | 6 | 810 |
| **Redmond<sup>16</sup>** | GSE151803 | Organoids | Pancreas | SARS-CoV-2 infected > mock treated | 3 | 3 | 393 |
| **Svendsen<sup>19</sup>** | GSE150392 | Human induced pluripotent stem cell-derived cells | Cardiomyocytes | SARS-CoV-2 infected > mock treated | 3 | 3 | 716 |
| **Takayama<sup>14</sup>** | GSE150819 | Organoids | Bronchial epithelial cells | SARS-CoV-2 infected > mock treated | 3 | 3 | 719 |
| **tenOever<sup>20</sup>** | GSE147507 | Cultured cells infected with SARS-CoV-2 | A549 | SARS-CoV-2 infected > mock treated | 3 | 3 | 480 |
| **tenOever<sup>20</sup>** | GSE147507 | Autopsy samples COVID patients | Lung | COVID > healthy | 2 | 2 | 576 |
| **tenOever<sup>20</sup>** | GSE147507 & GSE200074 | Autopsy samples COVID patients | Lung | COVID > healthy | 2 | 2 | 711 |
| **Ting<sup>15</sup>** | GSE150316 | Autopsy samples from COVID patients | Lung | COVID > healthy | 52 | 5 | 384 |

### Electronic health record analyses

The study was approved by the University of California, San Francisco, institutional review board. Data from the Cerner Real World Data COVID-19 deidentified EHR database containing records of 490,373 patients with a diagnosis of COVID-19 or COVID-19 exposure across 87 health care centers were analyzed. The following statins were included: atorvastatin, fluvastatin, lovastatin, pitavastatin, pravastatin, rosuvastatin, and simvastatin. Primary outcome was death after the onset of COVID-19. Inclusion criteria, considered comorbidities, and statistical analysis are detailed in the **Supplemental Methods**.

### Viral infection of Vero6 cells with SARS-CoV-2 virus

All work with native SARS-CoV-2 virus was performed in a BSL3 laboratory and approved by our Institutional Biosafety Committee. All drug screens to assess SARS-CoV-2 inhibition and cytotoxicity were performed with Vero E6 (Vero6) cells (ATCC# CRL 1586) using published methods (**Supplemental Methods**).^26^ A curve fitting procedure was used to determine IC50 and CC50 values (**Supplemental Methods**).

### Viral infection & host response of HUVECs with OC43 virus

To measure the impact of selected drugs on HCoV-OC43 infection, 96-well plates seeded with human umbilical vein endothelial cells (HUVECs) were infected with HCoV-OC43 and treated with drugs (**Supplemental Methods**). Viral load, Hoechst fluorescence, and IP-10 measurements were measured and normalized to vehicle control samples for each assay. Each group was compared to vehicle controls using the Brown-Forsythe and Welch ANOVA tests and corrected for multiple comparisons using a Dunnett T3 test.

### Visualizations

Plotting was performed in Prism 9 (GraphPad Software LLC) or in R versions 3.0.2 and 4.0.5. Schematic in Figure 1 was made in Biorender.

### Role of the funding source

Funding sources were not involved in study design, in the collection, analysis, and interpretation of data, or in the writing of the report.

## Results

The NeMoCAD gene network analysis tool^16^ was used to identify FDA-approved drugs predicted to normalize the COVID-19 gene expression profile based on transcriptomic signatures of human cells or organoids infected with SARS-CoV-2 as well as cells or tissues obtained from COVID-19 patients or healthy control subjects. NeMoCAD identified gene changes across the transcriptome, compared them with gene expression changes induced by approved drugs in existing databases (e.g., LINCS, KEGG, TRRUST, CTD), and then prioritized compounds based on their ability to shift the disease transcriptomic signature state back to a healthy state (**Figure 1A**). COVID-19 normalizing drugs were predicted based on 14 differential RNA-seq expression datasets (COVID-19 vs. healthy) from 12 independent transcriptomics studies (**Table 1**). Across all datasets, NeMoCAD prioritized a different number of drugs for each dataset (**Table 1**), with 172 drugs representing the intersection of all these results and therefore shared drugs relevant to all samples (**Table 2**). Of the 2,436 drugs we investigated across the 14 differential expression datasets, 1,477 of the 2,436 drugs were not predicted to normalize any disease signature. Across the 959 compounds predicted to normalize at least one disease signature, each drug was predicted on average by 8.1 of the 14 datasets. Surprisingly, we found multiple statins that inhibit HMGCR to be predicted more frequently than expected by the average, with simvastatin predicted 14/14 times, pravastatin 13/14 times, and lovastatin 12/14 times (**Figure 2A**). Of the 9 statins included in the NIH LINCS program database, 8 were in the top 25% of drugs predicted for at least one dataset investigated (**Figure 2B**). Across all datasets, simvastatin and fluvastatin were most frequently among the top 25% of predicted compounds.

**Table 2.**
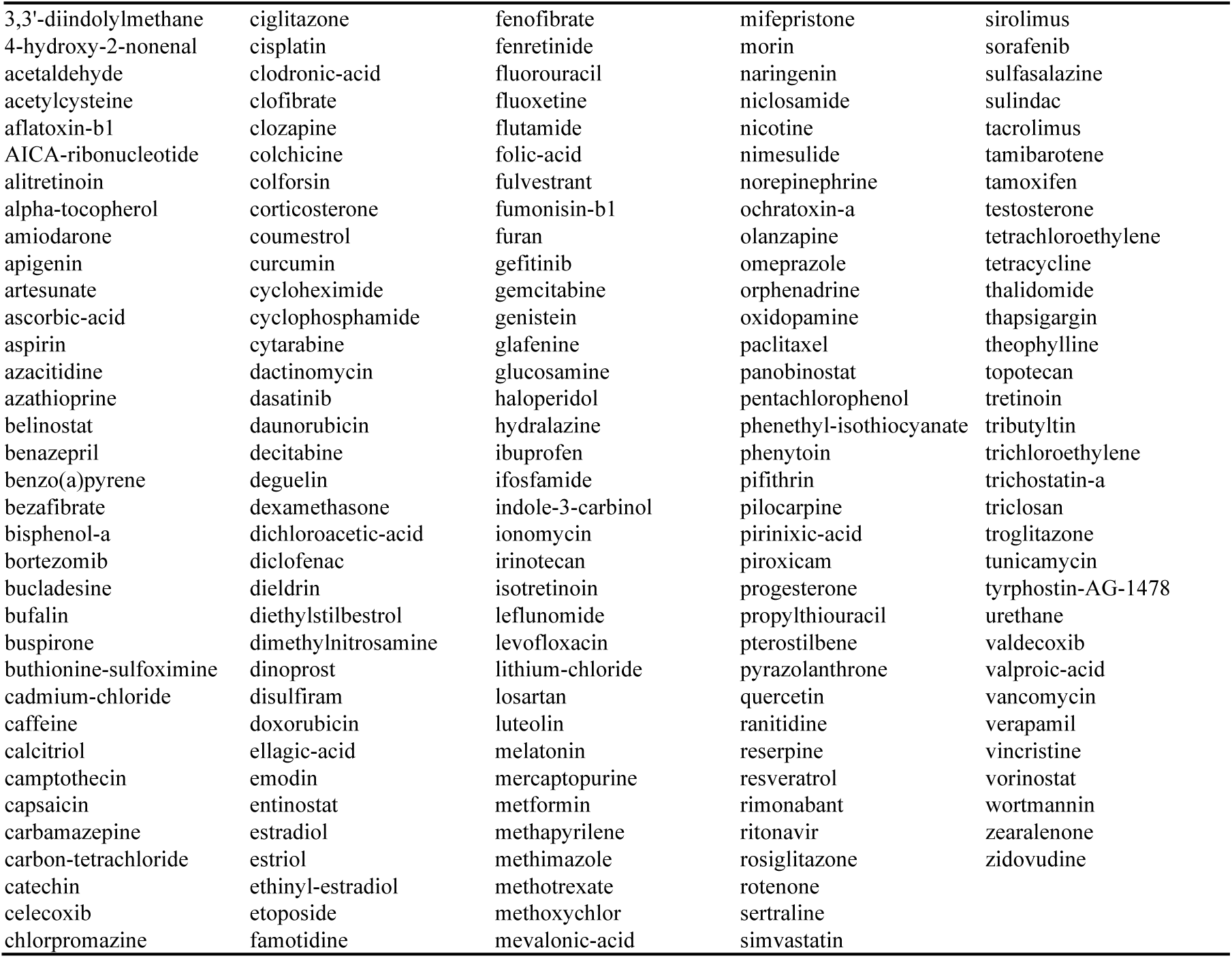
Drugs predicted 14 times out of the 14 state changes investigated.

**Figure 2.**
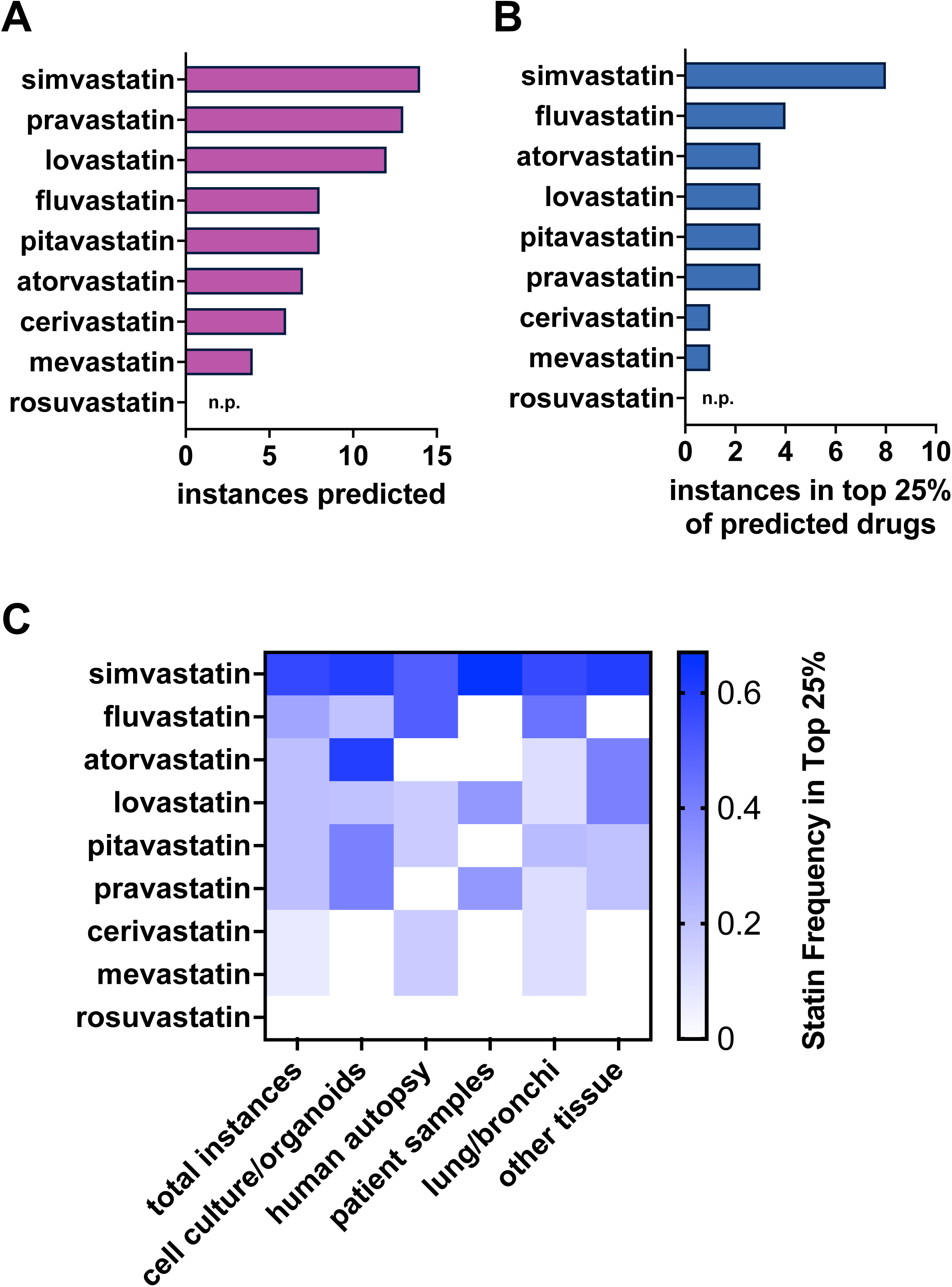
Simvastatin is identified by gene network-based predictions as the most likely drug in its category to reverse COVID-19 transcriptional profiles and potently inhibits SARS-CoV-2 *in vitro*. **(a)** Statins are predicted to shift the COVID-19 state to a healthy state, with simvastatin predicted for all datasets analyzed (14). Rosuvastatin was the only statin not predicted (n.p.) for any COVID-19 transcriptomics signatures. **(b)** 8 of 9 statins in the LINCS database were in the top 25% of drugs predicted for at least one dataset investigated. **(c)** Frequency of prediction for each statin when input datasets are stratified by sample source and tissue origin. Prediction frequency is normalized by the number of input datasets from each sample source and tissue origin.

We further assessed our predictions to understand how different types of input data might impact the types of compounds predicted. Stratification of the input datasets by sample source (COVID patient, autopsy sample, or cell culture/organoid) and tissue origin (lung/bronchi or other) revealed that simvastatin is frequently predicted across all dataset types (**Figure 2C**). In addition, atorvastatin is often predicted when cell culture and organoid samples are used as data inputs, whereas fluvastatin is commonly predicted in human autopsy samples, and lovastatin and pravastatin are predicted at an intermediate frequency using patient input data. Specific investigation of tissue origin revealed that simvastatin and fluvastatin are most frequently predicted when input datasets are derived from lung or bronchi tissue (**Figure 2C**). Simvastatin, atorvastatin, and lovastatin are also frequently predicted using samples from other non-lung tissues, including nasopharyngeal swabs, blood, liver, pancreas, and cardiac cells.

This finding that statins might differ in their ability to suppress responses to SARS-CoV-2 infection and that these effects could be independent of their common lipid lowering activity induced us to explore whether different statins also exhibit disparate activities in COVID-19 patients. We used the Cerner Real World Data COVID-19 deidentified EHR database to assess the effects of various statins on the survival of COVID-19 patients who were prescribed these medications. This large database represents a diverse population of patients diagnosed with COVID-19 from January to September 2020 with a duration of follow-up of as long as 8 months in 87 health centers across the US. Among 70,308 eligible patients, we identified 4,330 patients with who were prescribed atorvastatin, lovastatin, pravastatin, rosuvastatin, or simvastatin (**Figure 1B**). There were no patients who were prescribed fluvastatin in this database. The remaining 65,978 patients had no history of statin exposure (control patients). Cohort characteristics are shown in **Table 3** and **Table 4**. As disease severity could vary between the medication exposed and unexposed groups, we accounted for the type of encounter (Urgent care, ER, Admission for Observation, or Inpatient) at the time of COVID diagnosis and found that after matching, there was adequate balance in the encounter type between the compared medication exposed and unexposed groups, with the absolute value of standardized mean difference (SMD) of less than 0.1 for encounter type (**Supplemental Figures S1 and S2**).

**Table 3.**
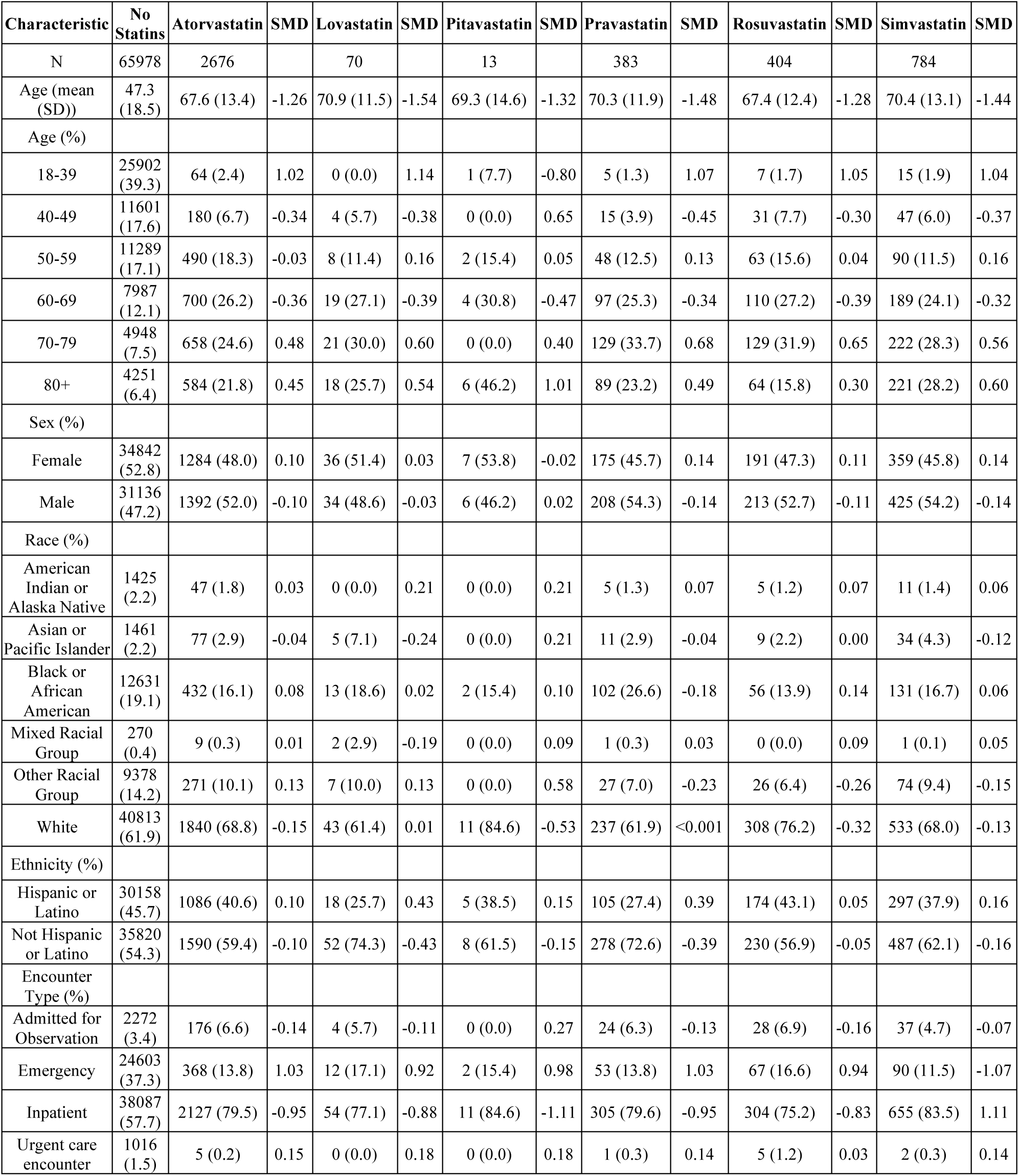
Cohort characteristics before propensity score matching (PSM), reflecting differing percentages of characteristics (demographics) with standardized mean differences (SMD) for those prescribed a specific statin compared to the control cohort not treated with a statin.

| Characteristic | No Statins | Atorvastatin | SMD | Lovastatin | SMD | Pitavastatin | SMD | Pravastatin | SMD | Rosuvastatin | SMD | Simvastatin | SMD |
| --- | --- | --- | --- | --- | --- | --- | --- | --- | --- | --- | --- | --- | --- |
| N | 65978 | 2676 |  | 70 |  | 13 |  | 383 |  | 404 |  | 784 |  |
| Age (mean (SD)) | 47.3 (18.5) | 67.6 (13.4) | -1.26 | 70.9 (11.5) | -1.54 | 69.3 (14.6) | -1.32 | 70.3 (11.9) | -1.48 | 67.4 (12.4) | -1.28 | 70.4 (13.1) | -1.44 |
| Age (%) |  |  |  |  |  |  |  |  |  |  |  |  |  |
| 18-39 | 25902 (39.3) | 64 (2.4) | 1.02 | 0 (0.0) | 1.14 | 1 (7.7) | -0.80 | 5 (1.3) | 1.07 | 7 (1.7) | 1.05 | 15 (1.9) | 1.04 |
| 40-49 | 11601 (17.6) | 180 (6.7) | -0.34 | 4 (5.7) | -0.38 | 0 (0.0) | 0.65 | 15 (3.9) | -0.45 | 31 (7.7) | -0.30 | 47 (6.0) | -0.37 |
| 50-59 | 11289 (17.1) | 490 (18.3) | -0.03 | 8 (11.4) | 0.16 | 2 (15.4) | 0.05 | 48 (12.5) | 0.13 | 63 (15.6) | 0.04 | 90 (11.5) | 0.16 |
| 60-69 | 7987 (12.1) | 700 (26.2) | -0.36 | 19 (27.1) | -0.39 | 4 (30.8) | -0.47 | 97 (25.3) | -0.34 | 110 (27.2) | -0.39 | 189 (24.1) | -0.32 |
| 70-79 | 4948 (7.5) | 658 (24.6) | 0.48 | 21 (30.0) | 0.60 | 0 (0.0) | 0.40 | 129 (33.7) | 0.68 | 129 (31.9) | 0.65 | 222 (28.3) | 0.56 |
| 80+ | 4251 (6.4) | 584 (21.8) | 0.45 | 18 (25.7) | 0.54 | 6 (46.2) | 1.01 | 89 (23.2) | 0.49 | 64 (15.8) | 0.30 | 221 (28.2) | 0.60 |
| Sex (%) |  |  |  |  |  |  |  |  |  |  |  |  |  |
| Female | 34842 (52.8) | 1284 (48.0) | 0.10 | 36 (51.4) | 0.03 | 7 (53.8) | -0.02 | 175 (45.7) | 0.14 | 191 (47.3) | 0.11 | 359 (45.8) | 0.14 |
| Male | 31136 (47.2) | 1392 (52.0) | -0.10 | 34 (48.6) | -0.03 | 6 (46.2) | 0.02 | 208 (54.3) | -0.14 | 213 (52.7) | -0.11 | 425 (54.2) | -0.14 |
| Race (%) |  |  |  |  |  |  |  |  |  |  |  |  |  |
| American Indian or Alaska Native | 1425 (2.2) | 47 (1.8) | 0.03 | 0 (0.0) | 0.21 | 0 (0.0) | 0.21 | 5 (1.3) | 0.07 | 5 (1.2) | 0.07 | 11 (1.4) | 0.06 |
| Asian or Pacific Islander | 1461 (2.2) | 77 (2.9) | -0.04 | 5 (7.1) | -0.24 | 0 (0.0) | 0.21 | 11 (2.9) | -0.04 | 9 (2.2) | 0.00 | 34 (4.3) | -0.12 |
| Black or African American | 12631 (19.1) | 432 (16.1) | 0.08 | 13 (18.6) | 0.02 | 2 (15.4) | 0.10 | 102 (26.6) | -0.18 | 56 (13.9) | 0.14 | 131 (16.7) | 0.06 |
| Mixed Racial Group | 270 (0.4) | 9 (0.3) | 0.01 | 2 (2.9) | -0.19 | 0 (0.0) | 0.09 | 1 (0.3) | 0.03 | 0 (0.0) | 0.09 | 1 (0.1) | 0.05 |
| Other Racial Group | 9378 (14.2) | 271 (10.1) | 0.13 | 7 (10.0) | 0.13 | 0 (0.0) | 0.58 | 27 (7.0) | -0.23 | 26 (6.4) | -0.26 | 74 (9.4) | -0.15 |
| White | 40813 (61.9) | 1840 (68.8) | -0.15 | 43 (61.4) | 0.01 | 11 (84.6) | -0.53 | 237 (61.9) | <0.001 | 308 (76.2) | -0.32 | 533 (68.0) | -0.13 |
| Ethnicity (%) |  |  |  |  |  |  |  |  |  |  |  |  |  |
| Hispanic or Latino | 30158 (45.7) | 1086 (40.6) | 0.10 | 18 (25.7) | 0.43 | 5 (38.5) | 0.15 | 105 (27.4) | 0.39 | 174 (43.1) | 0.05 | 297 (37.9) | 0.16 |
| Not Hispanic or Latino | 35820 (54.3) | 1590 (59.4) | -0.10 | 52 (74.3) | -0.43 | 8 (61.5) | -0.15 | 278 (72.6) | -0.39 | 230 (56.9) | -0.05 | 487 (62.1) | -0.16 |
| Encounter Type (%) |  |  |  |  |  |  |  |  |  |  |  |  |  |
| Admitted for Observation | 2272 (3.4) | 176 (6.6) | -0.14 | 4 (5.7) | -0.11 | 0 (0.0) | 0.27 | 24 (6.3) | -0.13 | 28 (6.9) | -0.16 | 37 (4.7) | -0.07 |
| Emergency | 24603 (37.3) | 368 (13.8) | 1.03 | 12 (17.1) | 0.92 | 2 (15.4) | 0.98 | 53 (13.8) | 1.03 | 67 (16.6) | 0.94 | 90 (11.5) | -1.07 |
| Inpatient | 38087 (57.7) | 2127 (79.5) | -0.95 | 54 (77.1) | -0.88 | 11 (84.6) | -1.11 | 305 (79.6) | -0.95 | 304 (75.2) | -0.83 | 655 (83.5) | 1.11 |
| Urgent care encounter | 1016 (1.5) | 5 (0.2) | 0.15 | 0 (0.0) | 0.18 | 0 (0.0) | 0.18 | 1 (0.3) | 0.14 | 5 (1.2) | 0.03 | 2 (0.3) | 0.14 |

**Table 4.** Cohort characteristics before propensity score matching (PSM), reflecting differing percentages of characteristics (conditions, and outcome of death) with standardized mean differences (SMD) for those prescribed a specific statin compared to the control cohort not treated with a statin.

| Characteristic | No Statins | Atorvastatin | SMD | Lovastatin | SMD | Pitavastatin | SMD | Pravastatin | SMD | Rosuvastatin | SMD | Simvastatin | SMD |
| --- | --- | --- | --- | --- | --- | --- | --- | --- | --- | --- | --- | --- | --- |
| N | 65978 | 2676 |  | 70 |  | 13 |  | 383 |  | 404 |  | 784 |  |
| Condition (%) |  |  |  |  |  |  |  |  |  |  |  |  |  |
| Obese | 23567 (35.7) | 1151 (43.0) | -0.15 | 24 (34.3) | 0.03 | 2 (15.4) | 0.48 | 157 (41.0) | -0.11 | 170 (42.1) | -0.13 | 298 (38.0) | -0.05 |
| Cancer | 2664 (4.0) | 188 (7.0) | -0.13 | 3 (4.3) | -0.01 | 1 (7.7) | -0.16 | 30 (7.8) | -0.16 | 25 (6.2) | -0.10 | 49 (6.2) | -0.10 |
| Cerebrovascular Disease | 2311 (3.5) | 378 (14.1) | 0.38 | 8 (11.4) | 0.31 | 2 (15.4) | 0.42 | 51 (13.3) | 0.36 | 60 (14.9) | 0.40 | 91 (11.6) | 0.31 |
| Chronic Kidney Disease (CKD) | 4650 (7.0) | 654 (24.4) | 0.49 | 10 (14.3) | 0.24 | 4 (30.8) | 0.64 | 121 (31.6) | 0.65 | 86 (21.3) | 0.42 | 178 (22.7) | 0.45 |
| COPD | 3460 (5.2) | 386 (14.4) | 0.31 | 7 (10.0) | 0.18 | 2 (15.4) | 0.34 | 60 (15.7) | 0.35 | 40 (9.9) | -0.18 | 113 (14.4) | 0.31 |
| Diabetes | 13677 (20.7) | 1586 (59.3) | -0.86 | 44 (62.9) | -0.95 | 8 (61.5) | -0.91 | 246 (64.2) | -0.98 | 232 (57.4) | -0.81 | 464 (59.2) | -0.85 |
| Heart Diseases | 7141 (10.8) | 1029 (38.5) | -0.68 | 22 (31.4) | -0.52 | 4 (30.8) | -0.51 | 154 (40.2) | -0.72 | 164 (40.6) | -0.73 | 280 (35.7) | -0.62 |
| Hypertension | 23146 (35.1) | 2206 (82.4) | -1.10 | 57 (81.4) | -1.07 | 11 (84.6) | -1.17 | 339 (88.5) | -1.32 | 337 (83.4) | -1.13 | 649 (82.8) | -1.11 |
| High Cholesterol | 9302 (14.1) | 1932 (72.2) | -1.45 | 57 (81.4) | -1.83 | 11 (84.6) | -1.99 | 304 (79.4) | -1.73 | 312 (77.2) | -1.64 | 592 (75.5) | -1.57 |
| Outcome (%) |  |  |  |  |  |  |  |  |  |  |  |  |  |
| Mortality | 4235 (6.4) | 431 (16.1) | 0.31 | 15 (21.4) | 0.44 | 4 (30.8) | 0.66 | 71 (18.5) | 0.37 | 53 (13.1) | 0.23 | 153 (19.5) | 0.40 |
| Healthcare Center Type (%) |  |  |  |  |  |  |  |  |  |  |  |  |  |
| Academic | 5689 (8.6) | 254 (9.5) | -0.03 | 4 (5.7) | 0.11 | 0 (0.0) | 0.43 | 46 (12.0) | 0.11 | 30 (7.4) | 0.04 | 60 (7.7) | 0.04 |
| Children | 263 (0.4) | 0 (0.0) | 0.09 | 0 (0.0) | 0.09 | 0 (0.0) | 0.09 | 0 (0.0) | 0.09 | 0 (0.0) | 0.09 | 0 (0.0) | 0.09 |
| Community Healthcare | 161 (0.2) | 6 (0.2) | 0.00 | 0 (0.0) | 0.07 | 0 (0.0) | 0.07 | 0 (0.0) | 0.07 | 0 (0.0) | 0.07 | 1 (0.1) | 0.03 |
| Community Hospital | 401 (0.6) | 13 (0.5) | 0.02 | 0 (0.0) | 0.11 | 0 (0.0) | 0.11 | 6 (1.6) | -0.09 | 3 (0.7) | -0.02 | 11 (1.4) | -0.08 |
| Critical Access | 39 (0.1) | 0 (0.0) | 0.03 | 0 (0.0) | 0.03 | 0 (0.0) | 0.03 | 0 (0.0) | 0.03 | 0 (0.0) | 0.03 | 0 (0.0) | 0.03 |
| IDN | 51485 (78.0) | 2108 (78.8) | -0.02 | 56 (80.0) | -0.05 | 10 (76.9) | 0.03 | 280 (73.1) | 0.12 | 309 (76.5) | 0.04 | 607 (77.4) | 0.02 |
| Regional Hospital | 7940 (12.0) | 295 (11.0) | 0.03 | 10 (14.3) | -0.07 | 3 (23.1) | -0.29 | 51 (13.3) | -0.04 | 62 (15.3) | -0.10 | 105 (13.4) | -0.04 |
| Region (%) |  |  |  |  |  |  |  |  |  |  |  |  |  |
| Northeast | 14216 (21.5) | 656 (24.5) | -0.07 | 11 (15.7) | 0.15 | 2 (15.4) | 0.16 | 61 (15.9) | 0.14 | 58 (14.4) | 0.19 | 162 (20.7) | 0.02 |
| Midwest | 5196 (7.9) | 207 (7.7) | 0.01 | 3 (4.3) | 0.15 | 0 (0.0) | 0.41 | 29 (7.6) | 0.01 | 24 (5.9) | 0.08 | 53 (6.8) | 0.04 |
| South | 26143 (39.6) | 1012 (37.8) | 0.04 | 23 (32.9) | 0.14 | 9 (69.2) | -0.62 | 194 (50.7) | -0.22 | 227 (56.2) | -0.34 | 289 (36.9) | 0.06 |
| West | 20423 (31.0) | 801 (29.9) | 0.02 | 33 (47.1) | -0.34 | 2 (15.4) | 0.38 | 99 (25.8) | 0.11 | 95 (23.5) | 0.17 | 280 (35.7) | -0.10 |

Overall, the propensity score distributions and SMDs of all matched covariates between treated and control groups before and after matching showed adequate balance between groups after matching, with absolute SMD values of less than 0.1 for all covariates, including demographics, heart diseases and other COVID-19 comorbidities, as well as conditions for which statins are prescribed (**Supplemental Figures S1 and S2**).

Importantly, among individual statins, we found that only treatment with atorvastatin, rosuvastatin, or simvastatin was associated with a statistically significant decrease in the relative risk of death in statin-treated patients compared to matched controls (**Table 5**). The mortality rate among atorvastatin-treated patients was 16.1% (431 of 2676) versus 20.4% (545 of 2676) among matched untreated control patients, with a reduction of 14% in the RR (0.86 [95% CI, 0.83-0.93]; Bonferroni adjusted p-value = 6.24E-05)(**Table 5 and Supplementary Table S1**). The mortality rate among rosuvastatin-treated patients was 13.1% (53 of 404) and 21.0% (850 of 4040) among matched untreated control patients, with a reduction of 41% in the RR (0.59 [95% CI, 0.45-0.78]; adjusted p-value = 9.61E-05)(**Table 5 and Supplementary Table S4**). The mortality rate among simvastatin-treated patients was 19.5% (153 of 784) and 23.3% (914 of 3920) among matched untreated control patients, with a reduction of 17% in the RR (0.83 [95% CI, 0.70-0,97]; adjusted p-value = 0.02)(**Table 5 and Supplementary Table S5**).

**Table 5.** Mortality rates of patients treated with (A) atorvastatin, (B) lovastatin, (C) pravastatin, (D) rosuvastatin, and (E) simvastatin, and matched control groups, and relative risk of death with 95% confidence interval and Benjamini-Hochberg adjusted p-value from the iteration with the least significant result for each comparison.

|  | treated patients |  | controls |  |  |  |
| --- | --- | --- | --- | --- | --- | --- |
| Statin (moderate dose) | Mortality rate, % | No. died/No. treated | Mortality rate, % | No. died/No. treated | Relative risk (95% CI) | Adjusted P-value* |
| Atorvastatin | 16.1 | 431/2676 | 20.4 | 545/2676 | **0.86**<br>**(0.80-0.93)** | 6.24E-05 |
| Lovastatin | 21.4 | 15/70 | 19.1 | 134/700 | 1.14<br>(0.66-1.96) | 0.76 |
| Pravastatin | 18.5 | 71/383 | 23.1 | 883/3830 | 0.78<br>(0.61-1.00) | 0.05 |
| Rosuvastatin | 13.1 | 53/404 | 21.0 | 850/4040 | **0.59**<br>**(0.45-0.78)** | 9.61E-05 |
| Simvastatin | 19.5 | 153/784 | 23.3 | 914/3920 | **0.83**<br>**(0.70-0.97)** | 0.02 |

Statins were also tested as part of a larger drug screening program in SARS-CoV-2-infected Vero6 cells. Within the statin drug class, simvastatin most potently inhibited infection with a half maximal inhibitory concentration (IC_50_) of 0.8 μM and almost a 10-fold higher 50% cytotoxic concentration (CC_50_ = 6.5 μM) (**Figure 3A-B**). The other statins were either unable to significantly inhibit SARS-CoV-2 infection in Vero6 cells or they were found to be toxic at doses required to see inhibitory effects (**Figure 3A and Supplemental Figure S3**). It is important to note that discordance observed between predictions made by NeMoCAD and SARS-CoV-2 inhibition in Vero6 cells is expected since NeMoCAD predicts drugs that will affect host response to infection, and not necessarily directly act on the virus to inhibit infection (e.g., reduce entry or replication). However, we also know that viral replication induces a host response and the transcriptional outcome of infection will always depend on interactions occurring within the virus-host system.^27,28^ Therefore, we cannot completely decouple the effects of predicted drugs on viral inhibition versus more conventional host measurements, such as cytokine levels. Furthermore, we found that simvastatin also inhibited infection of HUVECs by a related coronavirus (OC43) (**Figure 3C**) and potently reduced cytokine (IP-10) production without cytotoxic effects (**Supplemental Figure S4**). Similar results were observed with IL-6 and GM-CSF although the virus induced levels of these cytokines were variable. Thus, this particular statin appears to exhibit direct antiviral activity in addition to the HMGCR activity it shares with the other stains.

**Figure 3.**
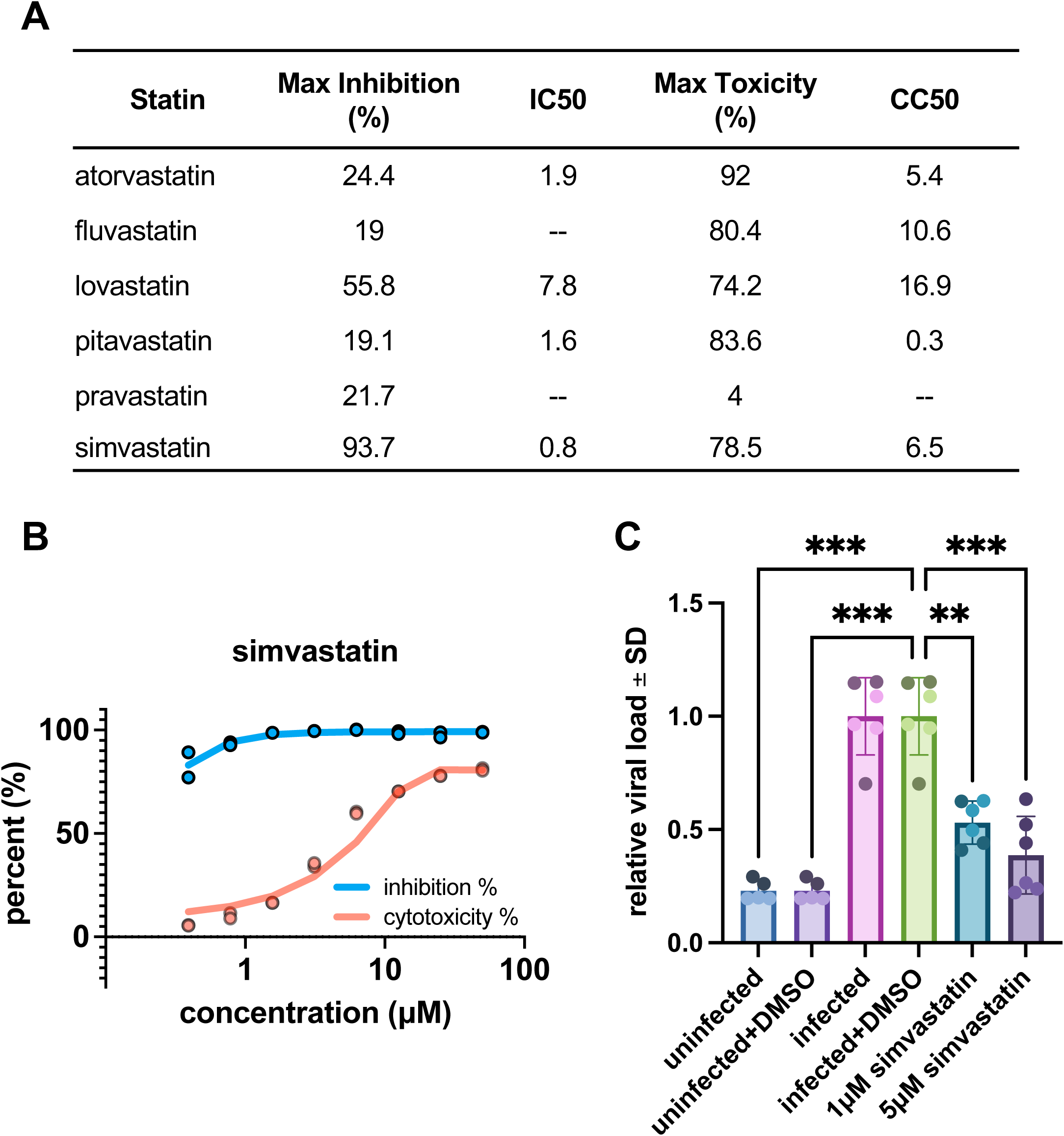
Effects of statins on coronavirus infections in vitro. **(a)** Inhibitory and cytotoxicity parameters from SARS-CoV-2 infection of Vero6 wild-type or GFP-expressing cells for a subset of statins contained in the LINCS database. Mean parameters for each statin are derived from two independent experiments. **(b)** Dose-response curves demonstrating the ability for simvastatin to inhibit GFP-SARS-CoV-2 infection (MOI=0.1) in a dose-dependent manner in Vero6 cells. **(c)** Simvastatin also inhibits the human coronavirus, OC43, in HUVEC cells when added at 1 μM or 5 μM concentrations (***p<0.001, **p<0.01). Error bars represent average +/- s.d.; repeated in n=2 independent biological experiments displayed in light (experiment 1) and dark (experiment 2) shaded data points.

## Discussion

Taken together, these data show that *in silico* prediction based on transcriptomics datasets from human patients, tissues, and cells combined with clinical database analyses offer a useful approach for identifying and validating non-obvious effects of FDA-approved drugs, thereby enabling rapid repurposing of these compounds. With multiple lines of evidence coming together, we found that clinical observations cannot always be explained by experimental biological studies, and vice versa. However, we demonstrate here that by looking at multiple lines of evidence in parallel, drug repurposing candidates are derisked across multiple biological levels to enable actionable repurposing interventions and potential new target discovery efforts.

Past retrospective studies suggested that patients prescribed drugs within the statin class have an overall lower risk of mortality from COVID-19,^4,6,8^ although this was not observed in one study.^10^ Predictions by NeMoCAD suggested that statins may differ in their ability to induce a shift from COVID-19 to healthy states, which could in part explain differences in results between these studies if the statin types differed.^4–6,10^ Specifically, NeMoCAD predicted that simvastatin, fluvastatin, and atorvastatin were the most likely statins to normalize the COVID-19 gene expression profile. Indeed, when we analyzed mortality in a large EHR database of patients with COVID-19, we confirmed that there are differences in the mortality risks of COVID-19 patients prescribed the different statins compared to their respective matched control cohort, with simvastatin, atorvastatin, and rosuvastatin associated with a significant reduction in the relative risk of death. We were unable to find a statistically significant difference in mortality risk among patients prescribed lovastatin or pravastatin represented in our EHR database compared to their respective matched control cohorts. Unfortunately, there were no patients who took fluvastatin and only 13 who took pravastatin in our EHR database, so we cannot make any conclusions about the protective effects of these compounds. Exploring an EHR database with a greater number of patients prescribed these statins in the future should allow for greater insights into any differences in mortality risk associated with these particular drugs.

Simvastatin and atorvastatin were predicted to be active by NeMoCAD, while rosuvastatin was not. Since the *in silico* predictions were based on the *in vitro* LINCS database, we expected some differences to be present between the predictions and EHR outcomes. NeMoCAD’s transcriptomics-based predictions would not take into account the physical properties (**Supplemental Table S6**) or potential antiviral activity for a drug. Structurally, rosuvastatin has a pyrimidine side ring, which makes it unique from the other statins which have pyrrole, naphthalene, or other side ring structures.^29^ Interestingly, remdesivir and molnupiravir, two of the few drugs to date that have received emergency use authorization (EUA) by the FDA for treatment of COVID-19,^30,31^ are nucleoside analogues -- chemically synthesized analogs of pyrimidines and purines.^32^ Remdesivir, molnupiravir, and other nucleosides have been shown to directly block SARS-CoV-2 infection in vitro and/or in animal models.^32–36^ The antiviral effects of nucleoside analogues is believed to result from their incorporation specifically by viral polymerases leading to defects in viral replication or through their antimetabolite activity where they compete with cellular enzymes for their natural ligands.^32^ We did not assess rosuvastatin in our in vitro screens; however, in work by Ahmed et al, their structure-based drug repositioning approach for drugs with potential inhibitory effects on Covid-19 virus predicted anti-viral drugs and rosuvastatin among their top six hits and demonstrated the inhibitory effects on SARS-CoV-2 replication by rosuvastatin and other drugs in VeroE6 cells.^37^ The physical properties of rosuvastin, in particular its structural similarity to nucleoside analogues, may confer this statin with additional antiviral properties not had by the other statins that we assessed, and could explain why our EHR analysis found rosuvastatin as associated with a reduction in the relative risk of death in patients with COVID-19 whereas NeMoCAD did not predict this drug.

Moreover, assessment of the LINCS data that defines the probability of each drug affecting specific genes combined with PCA also revealed closer clustering amongst simvastatin, lovastatin, and atorvastatin, which is consistent with their frequent co-prediction, whereas rosuvastatin is more distant (**Figure 4A**). Global analysis of the mortality-reducing statins also shows greater similarity in LINCS probability distribution between simvastatin and atorvastatin, whereas a wider probability distribution is observed for rosuvastatin (**Figure 4B)**, and a detailed comparison of the shared top gene targets (>90^th^ percentile) between these three statins similarly revealed that the most genes are shared between simvastatin and atorvastatin (**Figure 4C**). Therefore, these two statins may act in a more similar manner than rosuvastatin, which could influence the COVID-19 response through alternative mechanisms (e.g., direct antiviral and/or post-transcriptomic effects) that would not be detected using our transcriptomics-based computational approach. Additionally, various statins are typically dosed based on their cholesterol and lipid lowering properties, not based on their immune modulating properties; thus, a drug that appears to be most effective in silico for one of its “side-effect” properties might not appear to have a similar effect in vivo simply because of dosing decisions in practice. The discordance observed between drug predictions and EHR outcomes suggests that either technique alone is insufficient to identify drugs for clinical use and therefore the combination of methods is essential for narrowing the list of drug candidates before inclusion in randomized trials.

**Figure 4.**
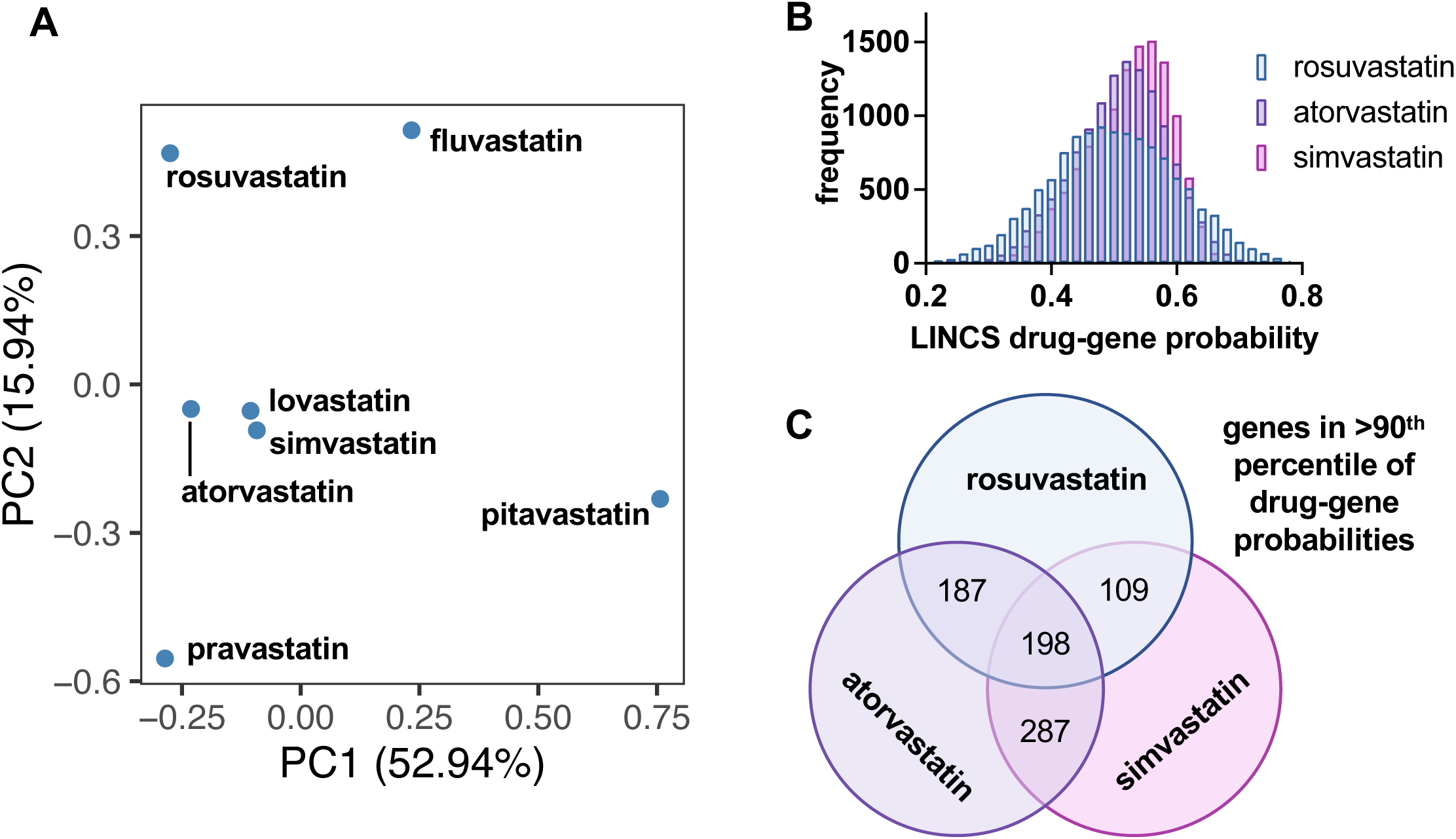
Comparison of LINCS drug-gene interaction probability data for each statin. **(a)** PCA plots of drug-gene interaction data for each statin reveals clustering of simvastatin, atorvastatin, and lovastatin when assessed across all genes in this dataset. **(b)** Distribution of LINCS drug-gene probabilities for statins that reduce mortality in patients. **(c)** Simvastatin, atorvastatin, and rosuvastatin share top gene targets (>90^th^ percentile), with the most shared between atorvastatin and simvastatin.

Collectively, our results suggest that statins exhibit divergent effects on the *host response* to SARS-CoV-2 infection despite a shared annotated target and common mechanism for treating dyslipidemia. In addition to their host-modulating effects, earlier work indicates that patients on statins seem to have improved outcomes following bacterial infection, an effect which is especially pronounced with respiratory tract infections, including pneumonia.^38^ However, meta-analysis of these studies reveals mixed results, again potentially suggesting that statins do not act uniformly as infection modulators. These protective effects of statins against infection may be due to their well-documented anti-inflammatory and immunomodulatory properties.^39,40^ Although originally developed to lower serum cholesterol, accumulating evidence suggests that statins have strong anti-inflammatory effects that contribute to their beneficial effects in patients experiencing vascular disease like atherosclerosis.^39^ Furthermore, statins may upregulate HO-1,^7^ which is a central modulator of the immune system, effecting anti-inflammation and anti-oxidation, which could prevent the severe “cytokine storm” inflammatory response that is central to morbidity and mortality in COVID-19 patients.^41^ By upregulating HO-1, statins, including simvastatin, lovastatin, atorvastatin, or rosuvastatin, also can increase the production of carbon monoxide and bilirubin,^9^ both of which have immunomodulatory, antioxidative and anti-inflammatory characteristics. In addition, statins may reduce the likelihood of graft-versus-host disease by inhibiting antigen presentation and shifting pro-inflammatory responses toward anti-inflammatory responses.^40^ We do not know precisely why certain statins appear to have superior disease-modifying activity than others. Unlike databases such as DrugBank that outline the known mechanisms of action, NeMoCAD surveys all possible interactions within the transcriptome and thus more work is needed to understand the clinical targets. However, longitudinal studies of the transcriptome in COVID-19 patients prescribed diverse statins could help clarify the mechanisms involved in these responses.

Despite strong evidence of their host modulating properties, there are also preliminary indications that some statins have more direct antibacterial and antiviral capabilities. Our *in vitro* studies with simvastatin suggest that at least this statin type can exhibit direct antiviral activity against both SARS-CoV-2 and the common cold coronavirus OC43, independently of its known lipid-lowering action. Simvastatin has previously been shown to exhibit superior antibacterial effects compared to fluvastatin and pravastatin, including against *S. pneumoniae* and *M. catarrhalis* infections.^38,42^ In the case of bacterial infection, it has been suggested that the activity of simvastatin is linked to its hydrophobicity, which may perturb the bacterial cell membrane compared to the more hydrophilic fluvastatin and pravastatin.^42^ Critically, the pro-drug form of simvastatin, predicted computationally and tested in our *in vitro* assays, is rapidly metabolized to its active acid form *in vivo* and therefore only a fraction of the total dose is maintained as the original pro-drug. Therefore, future work should investigate simvastatin combined with an inhibitor of cytochrome P450, a hemeprotein that plays a key role in the metabolism of drugs. By limiting drug metabolism, it would be possible to better assess if the simvastatin pro-drug is responsible for anti-viral effects *in vivo*.

Many of the potential COVID-19 therapeutics identified using past computational drug repurposing strategies failed when tested either using in vitro culture models, animals, or in the clinic. In fact, very preliminary *in vivo* studies by our team showed no effect of simvastatin on infection nor host response in SARS-CoV-2-infected hamsters and mice, despite attaining plasma levels that were greater than the IC_50_ for inhibition of infection and host response *in vitro*. These contrasting results highlight the challenges involved in conducting translational investigation from *in silico* predictions to *in vitro*, pre-clinical, and clinical studies, in particular raising questions about the clinical translation relevance of any disease model. Leveraging large patient databases early in the drug repurposing process to validate drugs predicted by computational approaches makes it possible to estimate how potential repurposed drugs may perform in infected patients and clinical sub-populations who regularly take these medications for other diseases or disorders. Importantly, in these databases we cannot completely rule out a possibility of unaccounted confounders correlated both with the drug and mortality, nor can we be confident that the medical histories of all patients are represented accurately as some information may be incomplete. Moreover, the retrospective nature of EHR analysis only allows us to identify an association between statin treatment and COVID-19 mortality, but not causal effects. Therefore, we envision that the process of generating *in silico* predictions and validating in databases will be a means to further narrow drug candidates and identify a more curated collection of therapeutics for testing in randomized control trials. Considering the continuing challenges with vaccine distribution and uptake, as well as the vulnerability of older populations to COVID-19, understanding non-obvious effects of approved drugs on patient mortality from infectious disease will be useful for combating this pandemic as well as ones that are likely to emerge in the future.

Finally, our findings suggest that drug repurposing efforts may require consideration of the many molecule-specific effects rather than taking reported drug targets and mechanisms at face value. Indeed, we believe that our approach, which counterpoints computational network-level analysis of biological interactions with *in vitro* exploratory screening and retrospective clinical evidence analysis, may form the basis of an altogether more powerful repurposing strategy in a pandemic scenario.

## Data Availability

All transcriptomics data used for drug predictions can be accessed through public databases indicated in Table 1. The Cerner patient data analyzed in this study is subject to the following licenses/restrictions: This was an observational study of electronic health records that cannot be made publicly available. Requests to access these datasets should be directed to the Cerner Clinical Research Team. All other data generated or analysed during this study are available from the corresponding author on reasonable request.

## Acknowledgements

The authors greatfully acknowledge funding from DARPA under Cooperative Agreements (grant nos W911NF-12-2-0036 to D.E.I. and W911NF-16-C-0050 to D.E.I. and M.F.) the Wyss Institute for Biologically Inspired Engineering at Harvard University (D.E.I.), the Christopher Hess Research Fund (D.K.S.) and in part by the University of California, San Francisco, Program for Breakthrough Biomedical Research grant (M.Si.), grant T32GM007618 from the Medical Scientist Training Program (M.Si.), and grant R35GM138353 from the National Institutes of Health (M.Si.). The content is solely the responsibility of the authors and does not necessarily represent the official views of the National Institutes of Health.

We thank Dr. B. tenOever for providing transcriptomics data; Dr. B. Oskotsky for discussions about and assistance with acquisition, analysis, and interpretation of data; and members of the Sirota Lab, University of California, San Francisco, for useful discussions. We also thank C. Akridge, G. Gasperino, K. Crane, and S. Purinton, along with everyone on the Cerner Clinical Research Team, for providing access to the Cerner Real World COVID-19 deidentified database and technical assistance; these individuals were not compenstated for their contributions.

## Declaration of interests

R.N. and D.E.I. hold equity in Unravel Biosciences, Inc and are members of its board of directors; D.E.I. is a member of its scientific advisory board; and R.N. is a current employee of the company. The remaining authors declare no competing interests.

## Contributors statements

R.N., M.Si., D.K.S. and D.E.I. conceived of and directed the work. M.M.S., S.K., and T.T. performed bioinformatic analysis and computational drug predictions with guidance from R.N. R.K.P., V.H., and K.E.C. oversaw the development and maintenance of the Wyss compound management database and COVID-19 transcriptomics database. M.R., B.F., M.So., P.P., J.L., and H.H. performed in vitro studies with guidance from G.G., K.E.C., and M.F. T.O., I.M., R.J.W., I.K., and B.L. developed analyses used to assess electronic health records with guidance by M.Si. and D.K.S. T.O. and I.M. performed the analysis of electronic health records. M.M.S., T.O., and D.E.I. drafted the manuscript and all authors reviewed and provided critical input for the manuscript. M.M.S. and T.O. verified the underlying data.

## Notes

### Author Declarations

The study was approved by the University of California, San Francisco, institutional review board.

### Summary of Updates

Title, clarifications in the Introduction and Results, new methods figure (Figure 1), additional Discussion added, and Supplemental files updated.

