## Supplement for "Target-agnostic drug prediction integrated with medical record analysis uncovers differential associations of statins with increased survival in COVID-19 patients"

**Supplemental Methods**

*NeMoCAD drug predictions*

Compounds were ranked based on correlation metrics (Pearson correlation and cross entropy) between the gene expression profiles of 2,436 compounds found in the LINCS database^1^ and target normalization signature. Independently, a network analysis approach using publicly available databases of gene-gene interactions (KEGG, TRRUST)^2–4^ and reference transcriptional signatures of drugs (LINCS, CTD)^1,5^ was used to calculate the probability of all drugs inducing the desired transcriptomic signature across all genes in the network. Drugs with a low probability score (probability<0.5) were filtered out from prediction lists. As a robustness criterion, drug predictions were assessed at multiple fold change thresholds to identify drugs that are consistently associated with the subnetwork defined by the differential expression profile. The frequency of NeMoCAD predictions across input datasets was evaluated for each statin within the LINCS database and statins with LINCS gene expression probabilistic network profiles that most strongly correlate with the target normalization signature were identified. We assessed the frequency of predictions within the top 25% of drugs, ranked by combination score (Pearson correlation and cross entropy) for each dataset. We selected the 25% threshold based on our prior experience using the NeMoCAD platform, during which we have typically selected predictions within the top quartile for further *in vitro* or *in vivo* screening. Prediction frequency was also stratified based on tissue source and type (**Table 1**).

*Human patient database*

This database represents patients with a diagnosis of COVID-19 or COVID-19 exposure who had an emergency department or urgent care visit or were admitted for observation or hospitalized. Subgroups of patients on a statin (atorvastatin, fluvastatin, lovastatin, pitavastatin, pravastatin, rosuvastatin, or simvastatin) exclude patients for whom more than 1 of the different statins was ordered during the inclusion period. Encounters include laboratory and pharmacy information (e.g., medication orders, dispensing), which are date and time stamped. Only patients with COVID-19 diagnosis confirmed by a laboratory test for SARS-CoV-2 (nucleic acid amplification tests and immunoassays) and/or by *International Statistical Classification of Diseases and Related Health Problems, Tenth Revision* (*ICD-10*) code U07.1 (for COVID-19 confirmed by laboratory testing),^6^ with known demographic characteristics (age, sex, and race and ethnicity), and who were 18 years or older from January to September 2020 were included in our study (n = 90,834). Patients with COVID-19, with a statin medication dose consistent with moderate-intensity therapy,^7^ and with a medication order status of active or completed and without a designation of as needed (ie, medication taken only when needed) at least once within a period of 10 days before and 7 days after their first recorded COVID-19 diagnosis were compared with patients with COVID-19 and no statin orders. We excluded patients with statin doses above or below those consistent with moderate-intensity therapy, with other order status (eg, inactive, discontinued, or unknown), or who were prescribed more than one statin type during this period or prescribed statins outside this period (n = 20,526), resulting in a final inclusion of 70,308 patients.

Considered comorbidities, identified by using *International Classification of Diseases, Ninth Revision, Clinical Modification* (*ICD-9-CM*), and *International Statistical Classification of Diseases, Tenth Revision, Clinical Modification* (*ICD-10-CM*), diagnosis codes, included hypertension (I10-I16 and 401.X-405.X), diabetes (O24, E11, E10, E13, and 250.X), chronic obstructive pulmonary disease (J44, 491.2, 493.2, and 496.X), obesity (E66.0, E66.1, E66.2, E66.8, E66.9, 278.00, 278.01, 278.03, Z68.3, Z68.4, V85.3, and V85.4), heart conditions (I20.X, I21.X, I22.X, I23.X, I24.X, I25.X, I42.X, I50.X, 410.X, 411.X, 412.X, 413.X, 414.X, 428.X, and 425.X), cerebrovascular disease (I6X.X, G45.X, G46.X, and 43X.X), cancer (C00-C96, 14X.X, 15X.X, 16X.X, 17X.X, 18X.X, 19X.X, and 20X.X), and chronic kidney disease (N18.X and 585.X). Prescription indications, identified by using *ICD-9-CM* and *ICD-10-CM* codes, included high cholesterol/hyperlipidemia (E78.0, E78.1, E78.2, E78.3, E78.4, E78.5, 272, 277). Body mass index (calculated as weight in kilograms divided by height in meters squared) values were identified by Logical Observation Identifiers Names and Codes 39156-5 and 89270-3. Body mass index values of at least 30 were used to confirm the diagnosis of obesity by *ICD-9-CM* or *ICD-10-CM* code. The mean dose prescribed for statins-treated patients was calculated for each statin. For patients receiving atorvastatin, the mean (SD) dose was 16.21 (4.85) mg/d. For patients receiving lovastatin, the mean (SD) dose was 40 (0) mg/d. For patients receiving pravastatin, the mean (SD) dose was 48.25 (16.21) mg/d. For patients receiving rosuvastatin, the mean (SD) dose was 8.65 (2.22) mg/d. For patients receiving simvastatin, the mean (SD) dose was 27.76 (9.74) mg/d.

*Statistical analysis*

The R Matchit package, version 3.0.2 (R Program for Statistical Computing) was used to perform propensity score matching with a nearest-neighbor method to match patients treated with a statin with control patients (1:1 ratio), atorvastatin-treated patients with control patients (1:1 ratio), lovastatin-treated patients with control patients (1:10 ratio), pravastatin-treated patients with control patients (1:10 ratio), rosuvastatin-treated patients with control patients (1:10 ratio), and simvastatin-treated patients with control patients (1:5 ratio), with varying ratios for the different statins to have sufficient power for the comparisons and increase precision.The propensity score was estimated using logistic regression of the treatment based on demographic characteristics (age, sex, and race and ethnicity), the encounter type at the time of the first recorded COVID-19 diagnosis, comorbidities (hypertension, diabetes, chronic obstructive pulmonary disease, obesity, heart conditions, cerebrovascular disease, cancer, and chronic kidney disease), and statin prescription indications (high cholesterol or hyperlipidemia) with age at encounter as a continuous variable, and the remaining as categorical variables. To assess covariate balance, density plots of the distribution of propensity scores for the treated and control groups before and after matching were created, and standardized mean differences between cohorts before and after propensity score matching were calculated.

We performed the Welch 2-sample, 2-sided *t*-test for continuous variables and the Pearson χ^2^ test with Yates continuity correction for categorical variables to evaluate whether there was a significant difference when comparing the two groups. We performed 10 iterations and evaluated mortality rate. Relative risks with 95% confidence intervals (Cis) were calculated for each iteration. Each iteration included all statin-treated patients and a subset of control patients chosen by propensity score matching. No statin-treated patients were discarded. We compared patients treated with (A) atorvastatin, (B) lovastatin, (C) pravastatin, (D) rosuvastatin and (E) simvastatin with their matched control patients. Owing to the large number of control patients, several of these patients could have had the same propensity score value, and therefore some statin-treated patients could be matched to several more than the number of control patients specified by the ratio (1, 5, or 10) having the same propensity score. To explore this variability and show the robustness of the results, we performed bootstrapping,^8^ with 10 iterations for each comparison, by varying the control patients who had a tie in their propensity scores. A significance threshold of .05 was applied to Benjamini-Hochberg–corrected *p* values. We reported he least significant result for each comparison.

*Viral infection of Vero6 cells with SARS-CoV-2 WT virus*

Cells were plated in opaque 96 well plates one day prior to infection. Drugs were diluted from stock to 50 μM and an 8-point 1:2 or 1:3 dilution series was prepared in duplicate in Vero Media. Every compound dilution and control was normalized to contain the same concentration of the respective drug vehicle. Cells were pre-treated with drug for 2 hours (h) at 37°C (5% CO2) prior to infection with SARS-CoV-2 (WA-1 strain - BEI #NR-52281) at multiplicity of infection (MOI) 0.01. In addition to plates that were infected, parallel plates were left uninfected to monitor cytotoxicity of drug alone. All plates were incubated at 37°C (5% CO2) for 3 days before performing CellTiter-Glo (CTG) assays as per the manufacturer’s instruction (Promega, Madison, WI). Luminescence was read on a BioTek Synergy HTX plate reader (BioTek Instruments Inc., Winooski, VT) using the Gen5 software (v7.07, Biotek Instruments Inc., Winooski, VT). Atorvastatin and pitavastatin were screened by this assay, which used a wild type (WT) version of SARS-CoV-2. The remaining statins were screened by a similar assay that used a GFP-labeled version of SARS-Cov-2.

*Viral infection of Vero6 cells with SARS-CoV-2 GFP virus*

The remaining statins were screened by a similar assay that used a GFP-labeled version of SARS-CoV-2. Sample dilutions were prepared as described for SARS-CoV-2 (WT) processing, with Vero6 cells (ATCC# CRL 1586) plated in clear bottom, black 96-well titer plates (Greiner bio-one, Kremsmünster, Austria) as opposed to opaque plates. Cell plates were pre-treated with drug for 2 h at 37°C (5% CO2) prior to infection with diluted SARS-CoV-2 GFP (generously provided by Dr. Ralph S. Baric) for a final MOI of 0.1. Plates were then incubated at 37°C (5% CO2) for 48 h, followed by fixation with 4.0% paraformaldehyde, nuclear staining with Hoechst (Invitrogen, Carlsbad, CA), and data acquisition on a Celigo 5-channel Imaging Cytometer (Nexcelom Bioscience, Lawrence, MA). The percent of infected cells was determined for each well by quantifying GFP expression by manual gating using the Celigo software. Toxicity plates were also processed as described in the WT SARS-CoV-2 screening. Pravastatin, lovastatin, simvastatin, and fluvastatin were screened by the GFP SARS-CoV-2 assay.

*Curve fitting of inhibition and cytotoxicity data*

Inhibition curves were fit using a sigmoidal [agonist] vs response curve with a fixed minimum value of 0. Fitting was attempted using two methods: first, fitting with the top of the curve unrestricted. If this failed, a second method was used wherein the top of the curve was constrained to the maximum inhibition or to 100%, whichever was greater. Relative IC50 (rIC50) was estimated using this fitted line, as the concentration at which 50% of maximum percent inhibition was achieved. If fitting failed, rIC50 was reported as the concentration at which 50% inhibition was exceeded by both replicates. If there were no values crossing the 50% threshold, we reported the maximum compound concentration tested in the run. Cytotoxicity curves were fit using a non-parametric Local Regression (Loess) curve with a fixed minimum value of 0. Relative CC50 (rCC50) was estimated using this fit line, as the concentration at which 50% of maximum percent cytotoxicity was observed.

*HUVEC assays*

To measure the impact of selected drugs on HCoV-OC43 infection, 96-well plates seeded with Human Umbilical Vein Endothelial Cells (HUVEC) were infected with HCoV-OC43 and treated with drugs. Uninfected and vehicle treated cells (0.1% DMSO) are included as controls. HUVEC plates were pre-dosed with drugs overnight at 34°C at indicated concentrations (100 µl/well). The following day, HCoV-OC43 was added to the wells (100 µl/well) at an MOI of 0.2 and incubated for 3 h at 34°C. After incubation, the medium containing virus and drugs was aspirated, the wells were washed with PBS +/+, new media was added with fresh drugs (100µl/well), and the plates were incubated for 72 h at 34°C. The supernatant was collected and stored for cytokine analysis and the plates were fixed with 4% paraformaldehyde for Hoechst and anti-viral staining. Fixed HUVECs were permeabilized with 100 µl/well of 0.1% Triton X, 1% FBS in PBS+/+ for 10 minutes at room temperature (RT), washed once with PBS, incubated in 100 µl/well of 1:50 human FcR Blocking Reagent (Miltenyi) in staining buffer (1% FBS in PBS+/+) for 30 minutes at RT, and washed again with PBS. Next, they were incubated with 100 µl/well of 1:400 Anti-OC43 coronavirus primary antibody (EMD Millipore MAB9013) in staining buffer for 30 minutes at RT, washed five times with PBS, and then incubated with 100 µl/well of 1:500 donkey anti-mouse HRP secondary antibody (Jackson ImmunoResearch 715-036-151) and 1:2000 Hoechst 33342 (Life Technologies H3570) in staining buffer for 1 hour at RT. After five PBS washes, Hoechst fluorescence was read at 355 nm excitation and 450 nm emission using a Synergy H1 Spectrophotometer. Subsequently, viral load was analyzed using the ImmPACT DAB Peroxidase (HRP) Substrate kit (Vector Labs) according to the manufacturer’s instructions, and DAB absorbance was read at 465 nm.

**Supplementary Figures**

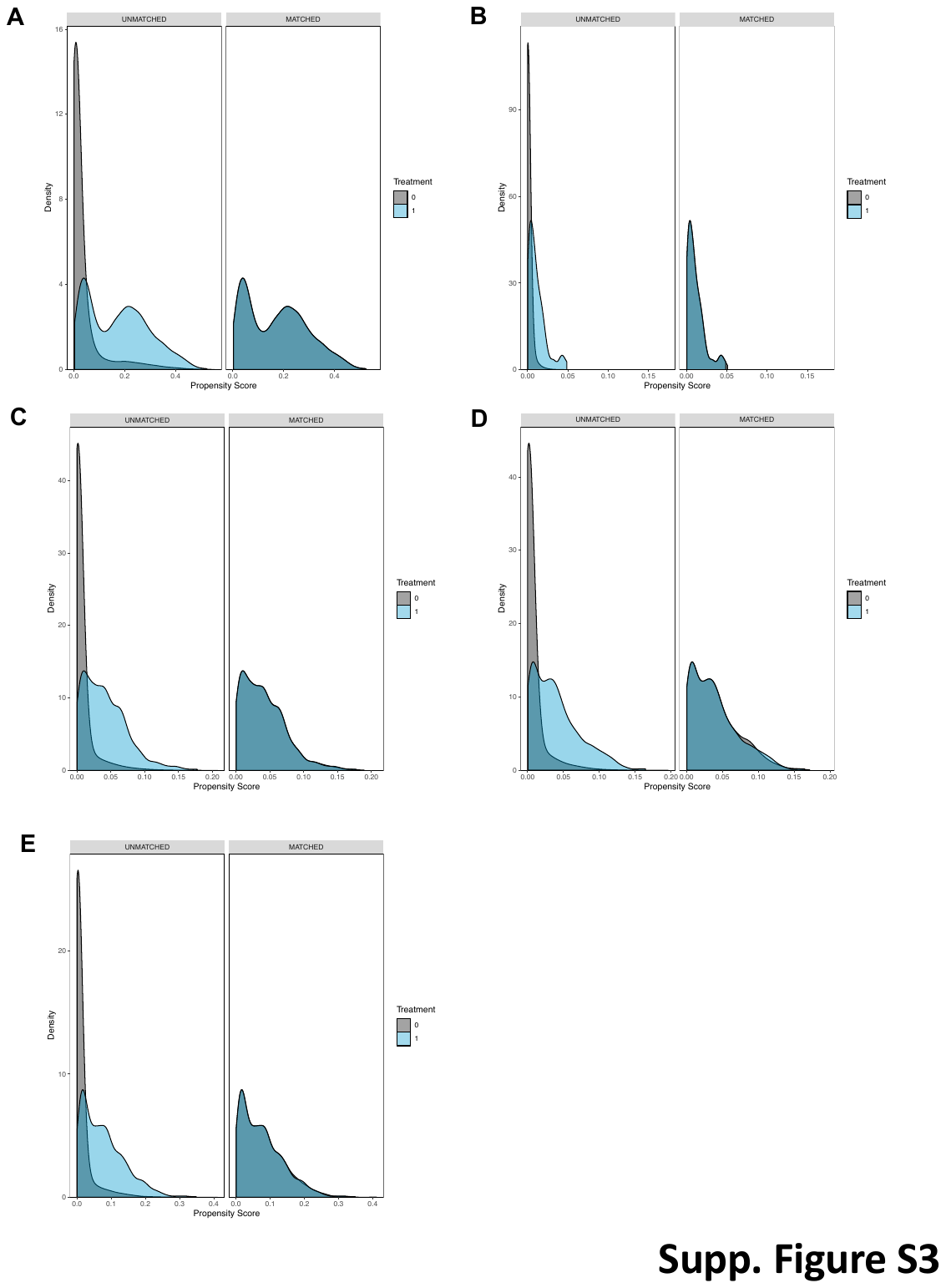

**Supplemental Figure S1.** Density plots of the distribution of propensity scores for the patients treated with (A) atorvastatin, (B) lovastatin, (C) pravastatin, (D) rosuvastatin, and (E) simvastatin, and control groups before and after matching.

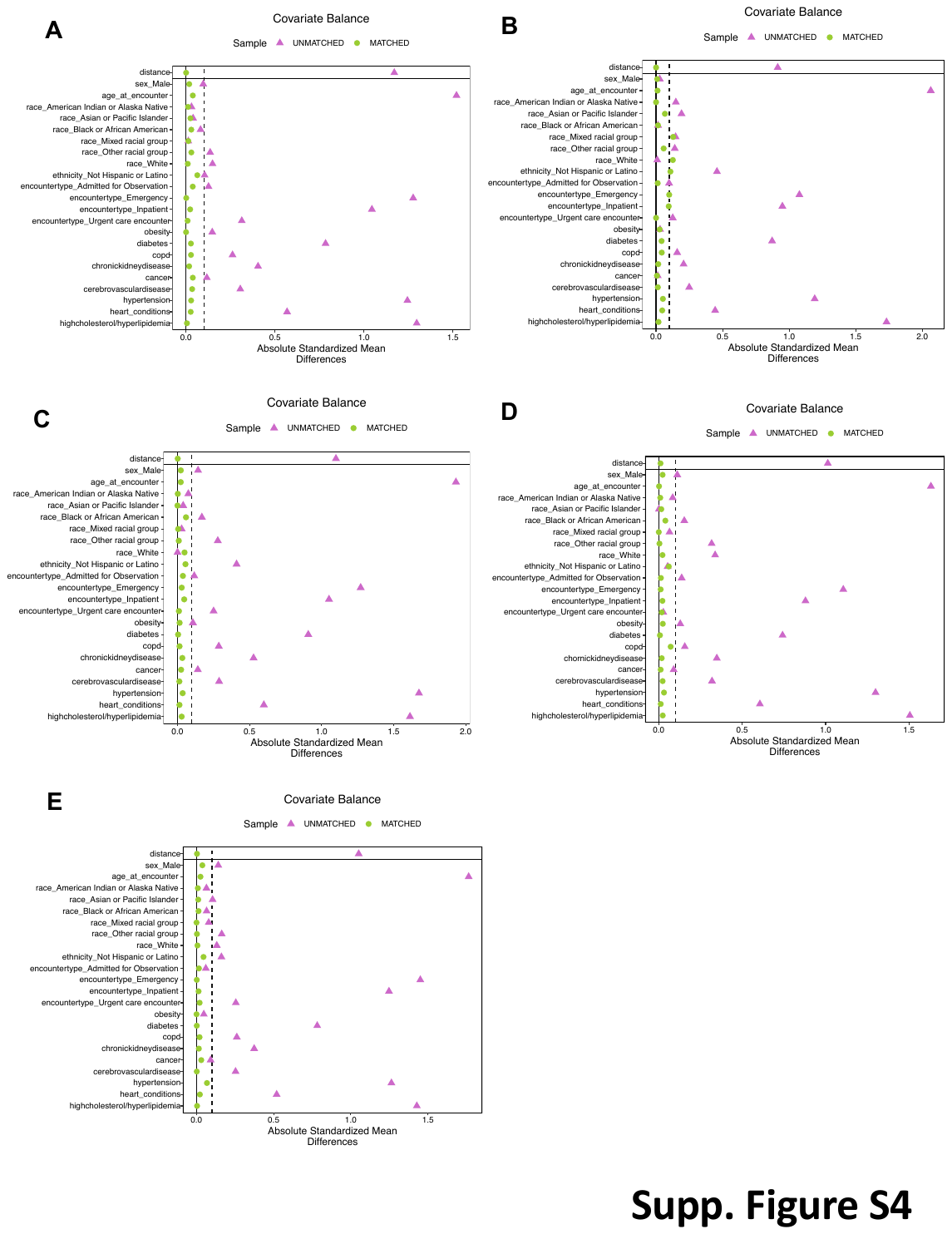

**Supplemental Figure S2.** Absolute standardized differences in means for matched categorical and continuous variables between patients treated with (A) atorvastatin, (B) lovastatin, (C) pravastatin, (D) rosuvastatin, and (E) simvastatin, and control groups for all data (before matching) and matched data.

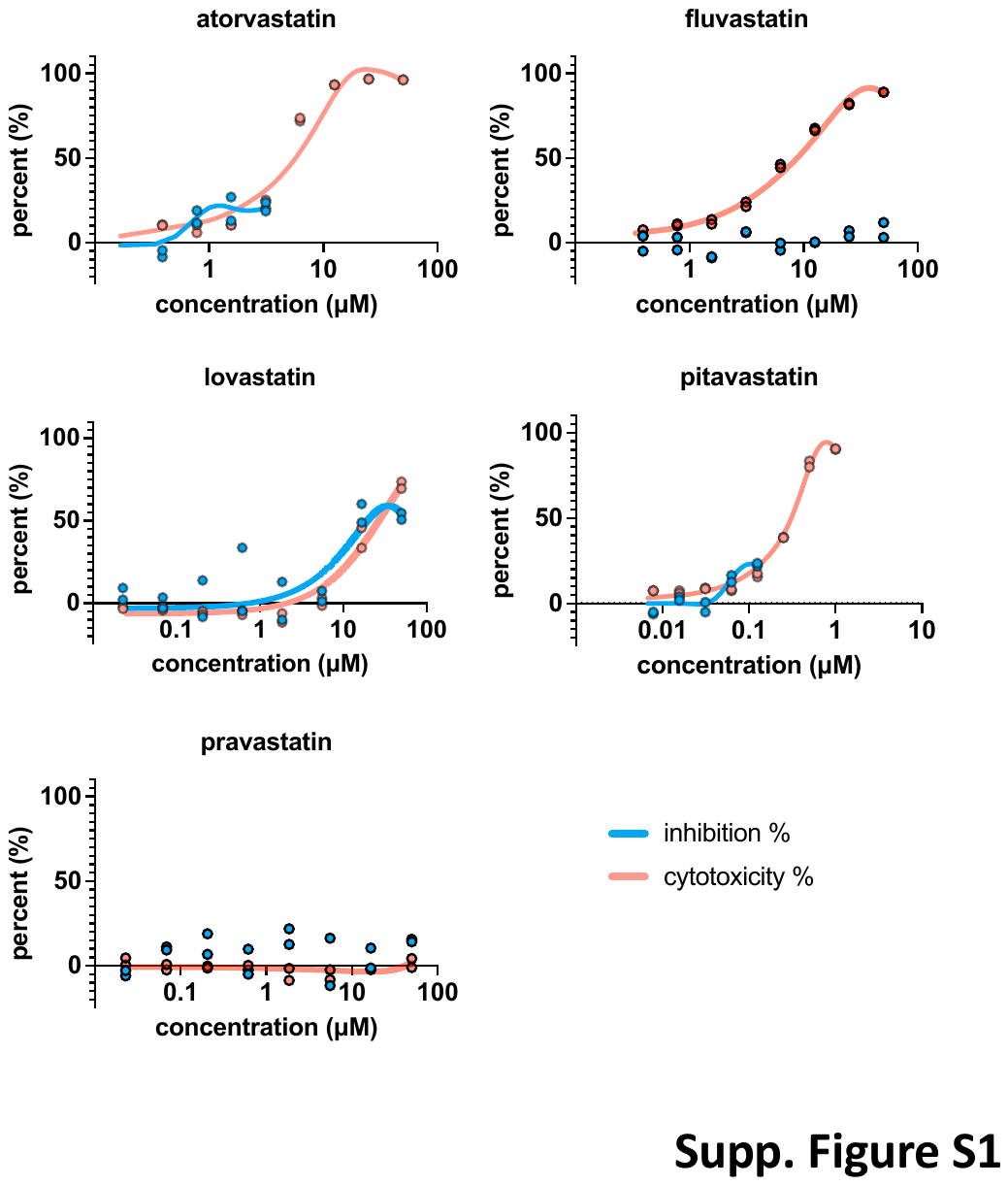

**Supplemental Figure S3.** Dose-response curves demonstrating the effects of statins on SARS-CoV-2 infection (MOI=0.1) in Vero6 cells.

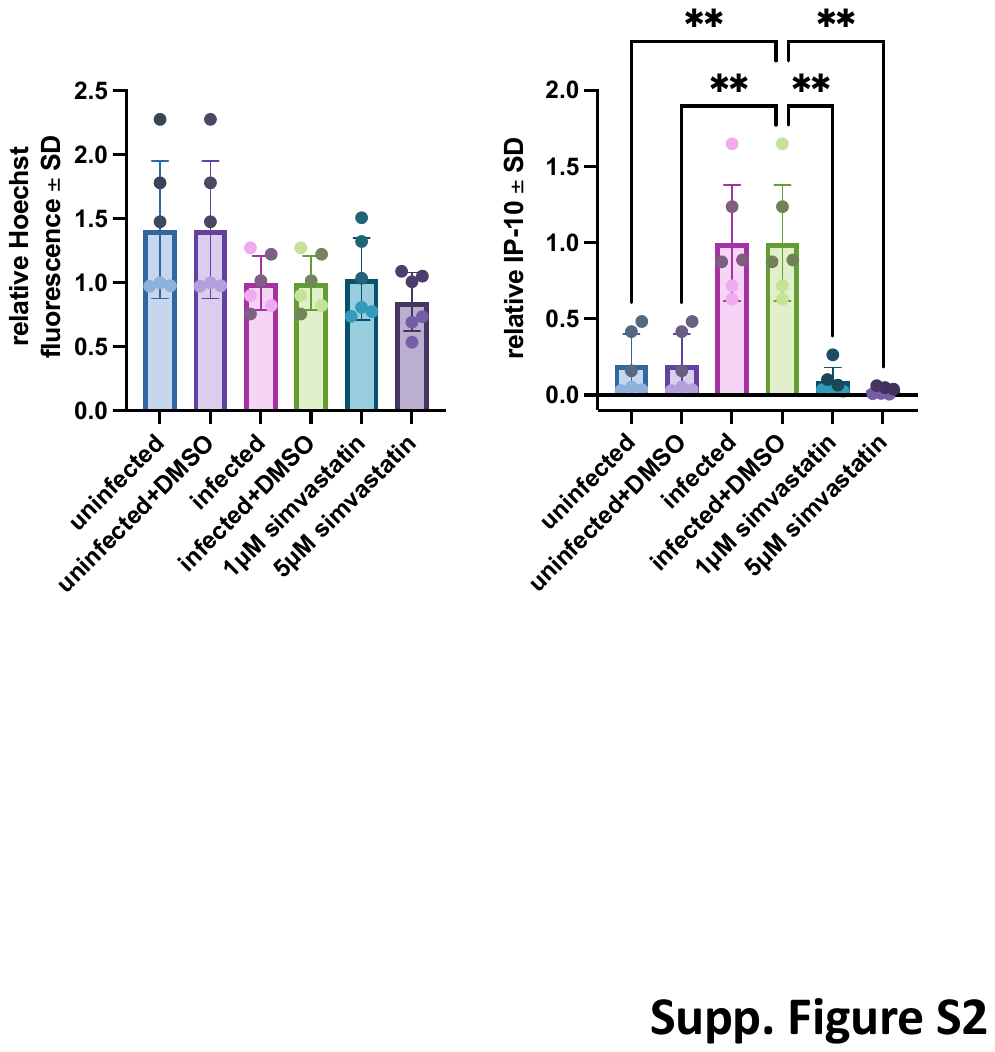

**Supplemental Figure S4.** Simvastatin added at 1μM or 5 μM concentrations in HUVEC cells infected with human coronavirus, OC43, modulates IP-10 levels without significant cytotoxicity detected by Hoechst fluorescence (***p<0.001, **p<0.01, *p<0.05). Error bars represent average +/- s.d.; repeated in n=2 independent biological experiments displayed in light (experiment 1) and dark (experiment 2) shaded data points.

**Supplementary Tables**

**Supplementary Table S1.** Propensity score (PS) matching by demographics, encounter type at the time of the first recorded COVID-19 diagnosis, COVID-19 comorbidities, and prescription indications: mortality rates for atorvastatin-exposed and PS-matched unexposed patients, a relative risk [95% confidence interval] and adjusted p-value in each iteration.

| **Iteration** | **Exposed to  atorvastatin, Died, N (%), (N total = 2676)** | **Matched controls, Died, N (%), (N total = 2676)** | **Difference in mortality rate (%)** | **RR [95% CI]** | **p-value (adjusted)** |
| --- | --- | --- | --- | --- | --- |
| 1 | 431 (16.1) | 557 (20.8) | -4.7 | 0.85 [0.79–0.92] | 2.60E-05 |
| 2 | 431 (16.1) | 564 (21.1) | -5.0 | 0.84 [0.78–0.91] | 1.71E-05 |
| 3 | 431 (16.1) | 553 (20.7) | -4.6 | 0.85 [0.79–0.92] | 2.74E-05 |
| 4 | 431 (16.1) | 568 (21.2) | -5.1 | 0.84 [0.78–0.90] | 1.71E-05 |
| 5 | 431 (16.1) | 545 (20.4) | -4.3 | 0.86 [0.80–0.93] | 6.24E-05 |
| 6 | 431 (16.1) | 550 (20.6) | -4.4 | 0.86 [0.79–0.92] | 3.76E-05 |
| 7 | 431 (16.1) | 554 (20.7) | -4.6 | 0.85 [0.79–0.92] | 2.74E-05 |
| 8 | 431 (16.1) | 555 (20.7) | -4.6 | 0.85 [0.79–0.92] | 2.74E-05 |
| 9 | 431 (16.1) | 561 (21.0) | -4.9 | 0.84 [0.78–0.91] | 1.85E-05 |
| 10 | 431 (16.1) | 548 (20.5) | -4.4 | 0.86 [0.80–0.93] | 4.49E-05 |

**Supplementary Table S2.** Propensity score (PS) matching by demographics, encounter type at the time of the first recorded COVID-19 diagnosis, COVID-19 comorbidities, and prescription indications: mortality rates for lovastatin-exposed and PS-matched unexposed patients, a relative risk [95% confidence interval] and adjusted p-value in each iteration.

| **Iteration** | **Exposed to  lovastatin, Died, N (%), (N total = 70)** | **Matched controls, Died, N (%), (N total = 700)** | **Difference in mortality rate (%)** | **RR [95% CI]** | **p-value (adjusted)** |
| --- | --- | --- | --- | --- | --- |
| 1 | 15 (21.4) | 138 (19.7) | 1.7 | 1.10 [0.64–1.89] | 0.757 |
| 2 | 15 (21.4) | 138 (19.7) | 1.7 | 1.10 [0.64–1.89] | 0.757 |
| 3 | 15 (21.4) | 137 (19.6) | 1.9 | 1.11 [0.64–1.91] | 0.757 |
| 4 | 15 (21.4) | 140 (20.0) | 1.4 | 1.08 [0.63–1.86] | 0.757 |
| 5 | 15 (21.4) | 138 (19.7) | 1.7 | 1.10 [0.64–1.89] | 0.757 |
| 6 | 15 (21.4) | 139 (19.9) | 1.6 | 1.09 [0.63–1.88] | 0.757 |
| 7 | 15 (21.4) | 139 (19.9) | 1.6 | 1.09 [0.63–1.88] | 0.757 |
| 8 | 15 (21.4) | 134 (19.1) | 2.3 | 1.14 [0.66–1.96] | 0.757 |
| 9 | 15 (21.4) | 137 (19.6) | 1.9 | 1.11 [0.64–1.91] | 0.757 |
| 10 | 15 (21.4) | 141 (20.1) | 1.3 | 1.07 [0.62–1.85] | 0.757 |

**Supplementary Table S3.** Propensity score (PS) matching by demographics, encounter type at the time of the first recorded COVID-19 diagnosis, COVID-19 comorbidities, and prescription indications: mortality rates for pravastatin-exposed and PS-matched unexposed patients, a relative risk [95% confidence interval] and adjusted p-value in each iteration.

| **Iteration** | **Exposed to  pravastatin, Died, N (%), (N total = 383)** | **Matched controls, Died, N (%), (N total = 3830)** | **Difference in mortality rate (%)** | **RR [95% CI]** | **p-value (adjusted)** |
| --- | --- | --- | --- | --- | --- |
| 1 | 71 (18.5) | 883 (23.1) | -4.5 | 0.78 [0.61–1.00] | 0.047 |
| 2 | 71 (18.5) | 889 (23.2) | -4.7 | 0.77 [0.60–0.99] | 0.047 |
| 3 | 71 (18.5) | 886 (23.1) | -4.6 | 0.77 [0.61–0.99] | 0.047 |
| 4 | 71 (18.5) | 883 (23.1) | -4.5 | 0.78 [0.61–1.00] | 0.047 |
| 5 | 71 (18.5) | 890 (23.2) | -4.7 | 0.77 [0.60–0.99] | 0.047 |
| 6 | 71 (18.5) | 883 (23.1) | -4.5 | 0.78 [0.61–1.00] | 0.047 |
| 7 | 71 (18.5) | 884 (23.1) | -4.5 | 0.78 [0.61–0.99] | 0.047 |
| 8 | 71 (18.5) | 883 (23.1) | -4.5 | 0.78 [0.61–1.00] | 0.047 |
| 9 | 71 (18.5) | 883 (23.1) | -4.5 | 0.78 [0.61–1.00] | 0.047 |
| 10 | 71 (18.5) | 886 (23.1) | -4.6 | 0.77 [0.61–0.99] | 0.047 |

**Supplementary Table S4.** Propensity score (PS) matching by demographics, encounter type at the time of the first recorded COVID-19 diagnosis, COVID-19 comorbidities, and prescription indications: mortality rates for rosuvastatin-exposed and PS-matched unexposed patients, a relative risk [95% confidence interval] and adjusted p-value in each iteration.

| **Iteration** | **Exposed to  rosuvastatin, Died, N (%), (N total = 404)** | **Matched controls, Died, N (%), (N total = 4040)** | **Difference in mortality rate (%)** | **RR [95% CI]** | **p-value (adjusted)** |
| --- | --- | --- | --- | --- | --- |
| 1 | 53 (13.1) | 857 (21.2) | -8.1 | 0.59 [0.44–0.78] | 9.61E-05 |
| 2 | 53 (13.1) | 855 (21.2) | -8.0 | 0.59 [0.45–0.78] | 9.61E-05 |
| 3 | 53 (13.1) | 855 (21.2) | -8.0 | 0.59 [0.45–0.78] | 9.61E-05 |
| 4 | 53 (13.1) | 850 (21.0) | -7.9 | 0.59 [0.45–0.78] | 9.61E-05 |
| 5 | 53 (13.1) | 854 (21.1) | -8.0 | 0.59 [0.45–0.78] | 9.61E-05 |
| 6 | 53 (13.1) | 860 (21.3) | -8.2 | 0.58 [0.44–0.77] | 9.61E-05 |
| 7 | 53 (13.1) | 854 (21.1) | -8.0 | 0.59 [0.45–0.78] | 9.61E-05 |
| 8 | 53 (13.1) | 856 (21.2) | -8.1 | 0.59 [0.44–0.78] | 9.61E-05 |
| 9 | 53 (13.1) | 853 (21.1) | -8.0 | 0.59 [0.45–0.78] | 9.61E-05 |
| 10 | 53 (13.1) | 854 (21.1) | -8.0 | 0.59 [0.45–0.78] | 9.61E-05 |

**Supplementary Table S5.** Propensity score (PS) matching by demographics, encounter type at the time of the first recorded COVID-19 diagnosis, COVID-19 comorbidities, and prescription indications: mortality rates for simvastatin-exposed and PS-matched unexposed patients, a relative risk [95% confidence interval] and adjusted p-value in each iteration.

| **Iteration** | **Exposed to  simvastatin, Died, N (%), (N total = 784)** | **Matched controls, Died, N (%), (N total = 3920)** | **Difference in mortality rate (%)** | **RR [95% CI]** | **p-value (adjusted)** |
| --- | --- | --- | --- | --- | --- |
| 1 | 153 (19.5) | 923 (23.5) | -4.0 | 0.82 [0.70–0.96] | 0.019 |
| 2 | 153 (19.5) | 918 (23.4) | -3.9 | 0.82 [0.70–0.97] | 0.019 |
| 3 | 153 (19.5) | 921 (23.5) | -4.0 | 0.82 [0.70–0.97] | 0.019 |
| 4 | 153 (19.5) | 921 (23.5) | -4.0 | 0.82 [0.70–0.97] | 0.019 |
| 5 | 153 (19.5) | 928 (23.7) | -4.2 | 0.81 [0.69–0.96] | 0.019 |
| 6 | 153 (19.5) | 924 (23.6) | -4.1 | 0.82 [0.69–0.96] | 0.019 |
| 7 | 153 (19.5) | 925 (23.6) | -4.1 | 0.82 [0.69–0.96] | 0.019 |
| 8 | 153 (19.5) | 923 (23.5) | -4.0 | 0.82 [0.70–0.96] | 0.019 |
| 9 | 153 (19.5) | 914 (23.3) | -3.8 | 0.83 [0.70–0.97] | 0.022 |
| 10 | 153 (19.5) | 919 (23.4) | -3.9 | 0.82 [0.70–0.97] | 0.019 |

**Supplementary Table S6.** Statin properties. Adapted from references 9 and 10.

| **Drug Name** | **Atorvastatin** | **Fluvastatin** | **Lovastatin** | **Pitavastatin** | **Pravastatin** | **Rosuvastatin** | **Simvastatin** |
| --- | --- | --- | --- | --- | --- | --- | --- |
| **Derivative** | Synthetic | Synthetic | Fungal | Synthetic | Fungal | Synthetic | Fungal |
| **Side Ring** | Pyrrole | Indole | Naphthalene | Quinoline | Naphthalene | Pyrimidine | Naphthalene |
| **Solubility** | Lipophilic | Lipophilic | Lipophilic | Lipophilic | Hydrophilic | Hydrophilic | Lipophilic |
| **Form Administered** | Active hydroxy acid | Active hydroxy acid | Inactive lactone | Active hydroxy acid | Active hydroxy acid | Active hydroxy acid | Inactive lactone |
| **Metabolism** | CYP3A4 | CYP2C9 | CYP3A4 | Non-CYP450 Limited CYP2C9/19 | Non-CYP450 | Non-CYP450 Limited CYP2C9/8 | CYP3A4 |
| **Clearance** | Hepatic | Hepatic | Hepatic | Hepatic | Hepatic and renal | Hepatic and renal | Hepatic |
| **Elimination half-life (hours)** | 15 to 30 | 0.5 to 2.3 | 2.9 | 12 | 1.3 to 2.8 | 19 | 2 to 3 |
| **Bioavailability (%)** | 12 | 19 to 29 | 5 | 51 | 18 | 20 | 5 |
| **Protein binding (%)** | 80 to 90 | >99 | >95 | 99 | 43 to 55 | 88 | 94 to 98 |
| **Active metabolites** | Yes | No | Yes | Yes | No | No | Yes |

5.	Lamb J, Crawford ED, Peck D, et al. *The Connectivity Map: Using Gene-Expression Signatures to Connect Small Molecules, Genes, and Disease*. www.broad.mit.edu/cmap.

6.	ICD-10 Version:2019. Published April 16, 2020. Accessed August 8, 2021. https://icd.who.int/browse10/2019/en#/U07.1

7.	Stone NJ, Robinson JG, Lichtenstein AH, et al. 2013 ACC/AHA Guideline on the Treatment of Blood Cholesterol to Reduce Atherosclerotic Cardiovascular Risk in Adults. *J Am Coll Cardiol*. 2014;63(25):2889-2934. doi:10.1016/j.jacc.2013.11.002

8.	Austin PC, Small DS. The use of bootstrapping when using propensity‐score matching without replacement: a simulation study. *Statistics in Medicine*. 2014;33(24):4306-4319. doi:10.1002/sim.6276

9.	Rosenson RS. Statins: Actions, side effects, and administration. In: *UpToDate*. Wolters Kluwer Health; 2021.

10.	Ward NC, Watts GF, Eckel RH. Statin Toxicity: Mechanistic Insights and Clinical Implications. *Circulation Research*. 2019;124(2):328-350. doi:10.1161/CIRCRESAHA.118.312782
